# Distinct Connectivity Signatures of Hallucinatory Experiences and Neuromelanin Signal in Adolescents

**DOI:** 10.64898/2026.05.10.26352847

**Authors:** Philip N. Tubiolo, Yash Patel, Alexandria Trepiccione, Katherine G. Jonas, Scott J. Moeller, Anissa Abi-Dargham, Roman Kotov, Jared X. Van Snellenberg, Greg Perlman

## Abstract

**Objective:** Late adolescence is a critical developmental period that typically precedes psychosis onset, yet the neural correlates of subclinical hallucinatory experiences that may impact psychosis risk are poorly understood. Given evidence from adult psychosis models implicating abnormal “triple network” connectivity among the frontoparietal (FPN), default mode (DMN) and salience/cingulo-opercular (CON) networks, as well as dopaminergic abnormalities, we examined whether hallucinatory experiences in adolescents are associated with altered triple network organization and dopamine-related measures in the midbrain.

**Methods:** We performed a cross-sectional analysis of 171 community adolescents aged 14-17 who underwent resting-state functional magnetic resonance imaging and neuromelanin-sensitive MRI. Hallucinatory experience severity was measured using the Specific Psychotic Experiences Questionnaire. Resting-state functional connectivity was calculated among *a priori* DMN, FPN, and CON cortical regions; we examined associations between connectivity, hallucinatory experience severity, within-network connectivity, system segregation, and neuromelanin signal in the ventral tegmental area (VTA).

**Results:** Greater hallucinatory experience severity was associated with stronger connectivity in a subnetwork composed of CON-DMN and CON-FPN edges. Greater hallucinatory experience severity was also associated with lower global network segregation. VTA neuromelanin signal was not directly associated with hallucinatory experience severity, but greater VTA signal predicted lower connectivity in the hallucination-related subnetwork. Greater VTA neuromelanin signal was also associated with a distinct pattern of stronger connectivity within DMN midline regions.

**Conclusions:** These findings implicate altered triple network organization in hallucinatory experiences during late adolescence and suggest that dopamine-related midbrain signal may reflect broader developmental variation in cortical network organization rather than symptom severity directly.

**Plain Language Summary:** Hallucinatory experiences during adolescence may signal increased risk for later psychotic disorders, but their brain basis is unclear. We studied 171 adolescents aged 14–17 using resting-state fMRI to measure brain network activity and neuromelanin-sensitive MRI to estimate dopamine-related midbrain signal. More severe hallucinatory experiences were linked to abnormal communication among three brain networks often implicated in psychosis. Dopamine-related signal was not directly related to hallucination severity but was associated with developmentally relevant network organization. Overall, this work serves to improve our understanding of the risk factors that may contribute to psychosis conversion in adulthood.

## 1. Introduction

Psychotic-like experiences (PLEs) are unusual thoughts and beliefs or perceptual disturbances that are common among the general population, particularly through childhood and adolescence where their prevalence is commonly reported to be between 10 and 20%^1,2^. Their persistence in childhood and adolescence is associated with delayed developmental milestones^3^, future PLEs in adulthood^4^, and a significantly heightened risk of psychosis conversion^5^. Perceptual abnormalities such as audiovisual hallucinatory experiences, while less commonly reported than other PLEs^6^, possess clinical significance that differs by age; while they may be a feature of typical development in early childhood^7^, persistence of attenuated hallucinatory experiences from late adolescence into early adulthood is associated with suicide attempts and lifetime diagnoses of psychotic and substance use disorders^8^. However, the neural correlates of hallucinatory experiences in late adolescence are poorly understood.

One hypothesis is that hallucinatory experiences in late adolescence share pathophysiological mechanisms with audiovisual hallucinations in adults with schizophrenia spectrum disorders, including dysconnectivity in the “triple network.” The triple network comprises the frontoparietal network (FPN, also referred to as the central executive network), default mode network (DMN), and cingulo-opercular, or salience, network (CON)^9^. While most classically implicated in cognitive and attentional deficits^10^, the triple network model of hallucinations posits that internally generated phenomena gain abnormal salience via processing from the CON (particularly the anterior insula^11^), resulting in or stemming from dysfunctional engagement with the FPN and DMN and misidentification of internal representations as externally sourced^10,12,13^. Prior work has identified specific abnormalities in resting-state functional connectivity (RSFC) within the triple network that are associated with positive and negative symptom severity in both schizophrenia and young adults experiencing attenuated psychosis^14,15^.

Emerging evidence suggests that this circuitry may already be abnormal in youth with PLEs, indicating that triple network dysfunction may be detectable before the typical age window of psychosis conversion. One cross-sectional study of children ages 9-11 in the Adolescent Brain Cognitive Development Study reported a positive association between DMN within-network RSFC and delusional ideation, and a negative association between CON within-network RSFC and perceptual distortions^16^. A separate 4-year longitudinal study demonstrated that adolescents with increasing PLEs across ages 12 to 16 displayed reduced DMN-CON RSFC, as well as increased RSFC between left and right insula when scanned in early adulthood^17^. A major gap in prior literature is how triple network RSFC is associated with PLEs in late adolescence, a particularly important developmental period immediately prior to the emergence of psychotic disorders.

A second unresolved question is whether dopaminergic signaling contributes to these large-scale network abnormalities in adolescence. The long-standing dopamine hypothesis of schizophrenia is supported by evidence from molecular imaging studies demonstrating dopaminergic disruption in individuals with psychotic disorders, particularly within the striatum^18^, as well as by the therapeutic benefit of medications that reduce dopaminergic function, and the induction of psychotic symptoms via pro-dopaminergic agents (e.g., amphetamine)^19,20^. However, direct evidence linking dopaminergic abnormalities to triple network disturbances implicated in hallucinatory experiences is limited, with prior work focused primarily on associations between salience-network connectivity and striatal dopamine measures^18,21^. Thus, beyond the well-established role of striatal dopamine dysfunction in psychosis, it is unclear whether developmental variation in midbrain dopaminergic systems is associated with cortical triple network connectivity. This question is especially relevant in adolescence, when mesolimbic and mesocortical pathways originating from the ventral tegmental area (VTA) continue to mature^22,23^ and refine their innervation of major regions of the triple network, including the lateral prefrontal cortex, anterior cingulate cortex, orbitofrontal cortex, anterior insula, hippocampus, ventral striatum, and amygdala^24^.

Addressing dopamine-related hypotheses in large pediatric samples requires a noninvasive approach capable of indexing dopamine-related signal in the midbrain. Given the practical limitations of studying dopaminergic function in these samples using invasive techniques such as positron emission tomography, neuromelanin-sensitive MRI (NM-MRI) has increasingly been employed as a safe and economical alternative for developmental research. NM-MRI indexes the concentration of NM in the substantia nigra (SN) and VTA, which accumulates in dopaminergic neurons via oxidation of cytosolic dopamine, and is thought to remain in cells unless and until cell death occurs^25^. Because the present study was designed to test associations with cortical triple network RSFC rather than striatal function per se, we focused *a priori* on the VTA, whose mesocorticolimbic projections are especially relevant to cortical and limbic targets central to the triple network. Recent studies suggest that psychotic disorders in adults are associated with increased neuromelanin-sensitive MRI (NM-MRI) signal in the SN-VTA^26,27^, and additional work has linked abnormal NM-MRI signal to lifetime substance use^28^ and chronic course of depression^29^, supporting its utility as a marker of dopaminergic variation. Despite this, few studies have utilized NM-MRI to characterize dopaminergic correlates of resting-state network organization in a developmental context, and none, to our knowledge, have used a multimodal imaging approach to investigate neural correlates of hallucinatory experiences in adolescents.

In the present study, we performed a multimodal investigation of triple network dynamics in 14–17-year-old adolescents. Participants underwent an imaging battery that included resting-state functional magnetic resonance imaging (rs-fMRI) and NM-MRI. To probe RSFC between cortical parcels defined *a priori* to comprise the DMN, FPN, and CON (which closely mirrors the salience network)^30^, we employed the network-based statistic (NBS)^31^, a graph-based technique that identifies connected subnetworks of suprathreshold edges associated with a variable of interest, such as hallucination experience severity. This approach is advantageous when effects are hypothesized to be distributed across interconnected circuits, rather than confined to isolated connections, because conventional edge-wise statistical testing would require correction across many individual connections and therefore reduce power to detect biologically meaningful network-level patterns. We hypothesized that hallucinatory experience severity would be associated with a connected subnetwork spanning the DMN, FPN, and CON, and that VTA NM-MRI signal would be related to connectivity within this subnetwork. By investigating whether triple network connectivity and dopamine-related midbrain signal jointly relate to hallucinatory experiences in late adolescence, this study addresses an important mechanistic gap during a key developmental period for psychosis risk.

## 2. Methods

### 2.1. Participants and Clinical Assessments

Data for this study comprise the baseline wave of the first 181 youth enrolled into a longitudinal study of adolescent brain development and mental health. Most participants (61.3%) identified as White non-Hispanic, and a majority (71.6%) of their parents had at least a bachelor’s degree. Participants were recruited using a commercial mailing list of homes with 14-to 17-year-olds, word of mouth, and community postings. Eligibility criteria included: fluency in English; ability to complete study procedures (e.g., read and answer surveys); and a legal guardian willing to attend the appointment, provide parental permission, and participate. Exclusion criteria included MRI contraindications (claustrophobia, implanted metal, etc.). Informed assent and consent were obtained from adolescents and parents, respectively, and the study was approved by the Stony Brook University Institutional Review Board.

Youth psychiatric symptoms and diagnoses were assessed using the Computerized Kiddie Schedule for Affective Disorders and Schizophrenia (KSADS-COMP)^32^. Binarized diagnostic groups were created based on youth report: Mood disorders (Major Depressive Disorder, Persistent Depressive Disorder, Bipolar Disorder I or II, or Disruptive Mood Dysregulation Disorder), anxiety disorders (Social Phobia, Specific Phobia, Panic Disorder, Agoraphobia, Generalized Anxiety Disorder), behavioral disorders (Attention Deficit Hyperactivity Disorder, Conduct Disorder, Oppositional Defiant Disorder), obsessive-compulsive disorder, post-traumatic stress disorder, eating disorder (Anorexia Nervosa, Bulimia Nervosa, Binge-Eating Disorder), and substance use disorder (Nicotine, Cannabis, Alcohol). Rates of disorders are presented in **Table 1**. Severity of psychotic-like experiences was assessed with the youth-reported Specific Psychotic Experiences Questionnaire (SPEQ)^33^. Given the specific relevance of hallucinatory experiences to this analysis, we focus on the hallucinatory experiences subscale, which comprises nine items taken from the Cardiff Anomalous Perceptions Scale^34^.

**Table 1.** Summary of participant demographic measures.

| **Participant Characteristic** |  |
| --- | --- |
| N | 171 |
| Imaging Data |  |
| Resting State fMRI (%) | 171 (100%) |
| NM-MRI (%) | 165 (96.49%) |
| Age, Years, mean (SD) | 16.22 (1.08) |
| Biological Sex, Female (%) | 69 (40.35%) |
| Ethnicity, Hispanic (%) | 36 (21.05%) |
| Race |  |
| African American (%) | 15 (8.77%) |
| Asian (%) | 10 (5.85%) |
| Caucasian (%) | 122 (71.35%) |
| More than one (%) | 19 (11.11%) |
| Other (%) | 5 (2.92%) |
| Stimulant Medication Use (%) | 22 (12.87%) |
| Reported Hallucinatory Experiences (%)<sup>a</sup> | 78 (45.61%) |
| SPEQ Hallucinations, mean (SD)<sup>b</sup> | 7.04 (7.01) |
| SPEQ Grandiosity, mean (SD) | 8.74 (5.15) |
| SPEQ Anhedonia, mean (SD) | 33.66 (8.49) |
| SPEQ Cognitive Disorganization, mean (SD) | 5.20 (3.16) |
| SPEQ Paranoia, mean (SD) | 13.29 (13.99) |
| N with Psychiatric Diagnosis (%)<sup>c</sup> | 105 (61.40%) |
| N with Multiple Diagnoses (%) | 69 (40.35%) |
| Any Mood Disorder (%) | 45 (26.32%) |
| Major Depressive Disorder (%) | 40 (23.39%) |
| Bipolar Disorder (%) | 8 (4.68%) |
| DMDD (%) | 1 (0.58%) |
| Any Anxiety Disorder (%) | 69 (40.35%) |
| Generalized Anxiety (%) | 24 (14.04%) |
| Panic Disorder (%) | 9 (5.26%) |
| Phobic Disorder (%) | 61 (35.67%) |
| Social Anxiety (%) | 27 (15.79%) |
| Any Behavioral Disorder (%) | 38 (22.22%) |
| ADHD (%) | 19 (11.11%) |
| Conduct Disorder (%) | 17 (9.94%) |
| ODD (%) | 20 (11.70%) |
| OCD (%) | 29 (16.96%) |
| Post-Traumatic Stress Disorder (%) | 14 (8.19%) |
| Eating Disorder (%) | 8 (4.67%) |
| Schizophrenia (%) | 1 (0.58%) |
| Substance Use Disorder (%) | 22 (12.87%) |

*Abbreviations:* fMRI, functional magnetic resonance imaging; SD, standard deviation; SPEQ, Specific Psychotic Experiences Questionnaire; DMDD, disruptive mood dysregulation disorder; ADHD, attention-deficit/hyperactivity disorder; ODD, oppositional defiant disorder; OCD, obsessive-compulsive disorder.
<sup>a</sup>A positive report of hallucinatory experiences was defined as any score above 0 on the SPEQ hallucination scale.
<sup>b</sup>SPEQ Hallucination mean and standard deviations across participants were calculated in participants with scores greater than zero, only.
<sup>c</sup>Psychiatric diagnoses were determined via the Kiddie Schedule for Affective Disorders and Schizophrenia.

For comparability with other studies of hallucinatory experiences in youth, we examined the association between hallucinatory experiences and KSADS-COMP diagnosis using Mann-Whitney U-tests. Additionally, differences in hallucinatory experience severity between participants with any diagnosis versus no diagnosis were assessed identically. False discovery rate was performed across all comparisons, a family of eight statistical tests. Effect sizes were calculated with Cliff’s delta.

### 2.2. MRI Acquisition

MRI acquisition was performed using a 3T Siemens (Erlangen, Germany) MAGNETOM Prisma Scanner with a Siemens 64-channel head-and-neck coil. After collecting T1-weighted and T2-weighted structural images with 0.8 mm^3^ isotropic voxels, as well as two spin echo field maps with alternating phase encoding directions, resting-state fMRI was acquired with the following parameters: TR = 800 ms, TE = 25 ms, 2.0 mm^3^ isotropic voxels, 204×204 mm field of view, multiband acceleration factor of 6, alternating anterior-posterior and posterior-anterior phase encoding directions, and 563 volumes collected per run. Neuromelanin-sensitive MRI was acquired using a 2D gradient recalled echo sequence with magnetization transfer contrast with TR=555 ms, TE=4.11 ms, flip angle=40°, magnetization transfer frequency offset = 1,200 Hz, 0.4 mm × 0.4 mm in-plane resolution, 1.5 mm slice thickness, 20 slices, and up to 11 total excitations (averaged offline to produce a single volume after excluding high-motion volumes; see *2.3 fMRI and Neuromelanin-Sensitive MRI Preprocessing*).

### 2.3. fMRI and Neuromelanin-Sensitive MRI Preprocessing

All structural and fMRI data underwent preprocessing through the Human Connectome Project Minimal Preprocessing Pipelines version 4.7.0^35^. Briefly, data was processed through the *PreFreesurfer, Freesurfer, PostFreesurfer, fMRIVolume,* and *fMRISurface* pipelines. Resting-state fMRI data was resampled to Connectivity Informatics Technology Initiative (CIFTI) format with 91,282 total greyordinates.

NM-MRI was processed as described elsewhere using a standard pipeline^28,29,36^. Each participant’s NM-MRI was visually inspected for acquisition-related and head motion artifact, with low-quality volumes being discarded. Notably, participants were fully excluded if their NM-MRI data comprised less than five volumes of sufficient quality. Mean NM-MRI signal for the bilateral VTA subregion was extracted for 29 voxels based on MNI coordinates reported by an independent group to represent the location of the VTA at a probability greater than 50%^37^.

While the VTA was identified *a priori* as our primary NM-MRI region based on its projections to cortical triple network regions, we also isolated a 249-voxel, bilateral SN pars compacta (SNc) region of interest (ROI) from the same subcortical atlas as VTA for use in supplementary analyses due to the established role of nigrostriatal dopaminergic dysfunction in the pathophysiology of schizophrenia^18^. Contrast-to-noise ratio in each of these ROIs was calculated using previously described methods^26^.

### 2.4. fMRI Regions of Interest

ROIs for analysis of resting-state fMRI were derived from the Cole-Anticevic Brain Network Parcellation (CABNP)^30^, which comprises 360 cortical and 358 subcortical parcels each assigned to one of twelve functionally defined, resting-state brain networks. For this study, only cortical parcels from each of the frontoparietal network (FPN), default mode network (DMN), and cingulo-opercular network (CON), an analogue the salience network, were considered, resulting in a total of 183 parcels and 16,654 possible connectivity edges.

### 2.5. Functional Connectivity Calculation

All functional connectivity processing procedures are described in the **Supplementary Methods**. ROI pair RSFC was calculated as the partial Pearson correlation between ROI timeseries, controlling for band-pass filtered motion parameters (MPs; using the same 0.009-0.08 Hz band-pass filter that was applied to each timeseries), their squares, derivatives, and squared derivatives, white matter signal and its derivative, cerebrospinal fluid signal and its derivative, and the global signal and its derivative. This calculation was repeated without controlling for global signal and its derivative (i.e., without global signal regression; GSR) for *post-hoc* sensitivity analyses, but all primary analyses employ GSR. All partial correlation values were Fisher’s r-to-Z transformed prior to further analysis.

### 2.6. Functional Connectivity Network Analysis

To determine RSFC subnetworks collectively associated with hallucinatory experience severity, we employed the network-based statistic (NBS)^31^, a permutation-based approach that controls the family-wise error rate at the level of connected clusters of suprathreshold edges, rather than edgewise control in a mass-univariate analysis. NBS was performed using the NBS Toolbox v1.2 with the following parameters: an initial T-statistic threshold of 3.1 (approximately equivalent to p = 0.002, two-tailed), 5,000 permutations, and a cluster extent approach. One-tailed tests were performed separately testing for positive and negative associations of functional connectivity with SPEQ hallucination score, respectively, controlling for age, sex at birth, and binary stimulant medication exposure status; Bonferroni correction was employed across contrasts. Nodes in resulting significant subnetworks were characterized by their degree centrality, or the number of edges formed in the network containing each node. Additionally, a robust multiple linear regression with Huber weighting was used to investigate associations between mean RSFC across all edges in the subnetwork and VTA NM-MRI signal, controlling for the same covariates as the NBS analysis. As a supplementary analysis, associations between mean RSFC and SNc NM-MRI signal were also assessed.

Given the exploration of associations between RSFC and both VTA NM signal and hallucinatory experience severity, we also investigated whether a direct association existed between these two variables, performed as a robust linear regression predicting hallucinatory experience severity from VTA NM controlling for the same covariates stated previously.

As an additional *post-hoc* analysis, we recalculated RSFC in the NBS-isolated edges without GSR to aid in the interpretation of the directionality of associations between RSFC and hallucinatory experience severity (as GSR induces RSFC anticorrelations as a consequence of removing shared global BOLD fluctuations^38^). Mean RSFC was then calculated separately for all edge classifications (DMN-CON, FPN-CON, and CON-CON), and across all edges collectively. Separate robust multiple linear regressions were performed to estimate the association between mean RSFC and SPEQ hallucination score.

Finally, we performed two additional NBS analyses to probe the specificity of observed hallucination-associated connectivity patterns. To demonstrate specificity with respect to other PLE dimensions, we repeated the primary NBS analysis described above while additionally covarying for SPEQ grandiosity, cognitive disorganization, anhedonia, and paranoia subscale scores. Next, to demonstrate that the hallucination-associated connectivity patterns could not be accounted for by overall burden of psychopathology, we repeated the primary NBS analysis covarying for the total number of diagnoses assigned according to the KSADS-COMP.

### 2.7. Within-Network Connectivity and System Segregation

To better characterize whole-network RSFC associations with hallucinatory experience severity, we first performed robust multiple linear regressions with Huber weighting predicting mean within-network RSFC from SPEQ hallucination score, controlling for age, sex at birth, and binary stimulant medication exposure status. Next, we calculated network-specific and across-network system segregation *S*, defined as 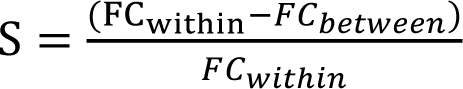, where 𝐹𝐶_𝑤𝑖𝑡ℎ𝑖𝑛_ is the mean of all within-network RSFC edges, either in a single network or across all three networks, and 𝐹𝐶_𝑏𝑒𝑡𝑤𝑒𝑒𝑛_ is the mean of all between-network RSFC edges^39^. False discovery rate correction was performed separately for the within-network connectivity (three tests) and system segregation (four tests) analyses at α = 0.05. This set of analyses was repeated replacing SPEQ hallucination score with VTA NM signal.

### 2.8. Exploratory Neuromelanin Network-Based Statistic Analysis

Given findings from primary analyses suggesting involvement of specific DMN nodes in the hallucination-associated subnetwork, as well as positive associations between VTA NM signal and DMN within-network RSFC strength, we performed an exploratory NBS analysis to spatially localize a connected subnetwork of edges whose RSFC was positively associated with VTA NM signal. This analysis was performed identically to the primary NBS analysis described above (see *2.6 Functional Connectivity Network Analysis*). Resulting nodes and edges were compared to the hallucination-specific subnetwork to identify areas of overlap between the two networks.

## 3. Results

### 3.1. Demographics

Demographic information for participants retained for analysis is shown in **Table 1**. After rigorous quality control evaluations and volume censoring, 171 participants were retained for analysis of resting-state fMRI, alone, of which 165 participants were retained for NM-MRI analyses. For *post-hoc* analyses of RSFC calculated without GSR, 159 participants were retained after volume censoring. Of the 171 total participants retained for primary analyses, 78 (45.61%) reported hallucinatory experience scores greater than zero (75, or 45.45%, in the NM-MRI sample of 165 participants).

A total of 105 participants (61.40%) were assigned at least one current or past psychiatric diagnosis, with 69 participants (40.35%) receiving multiple diagnoses. A comparison of hallucinatory experience severity by diagnosis is shown in **Figure S1**. Hallucinatory experience severity was greater in participants with mood disorders, anxiety disorders, and behavioral disorders (**Figure S1A-C**). Additionally, hallucinatory experience severity was greater in participants with at least one diagnosis across any category, relative to those with zero diagnoses (**Figure S1H**).

### 3.2. Functional Connectivity Network Analysis

NBS identified a connected subnetwork with 47 parcels among the cortical ROIs (see **Figure 1A**; 20 from CON, 21 from DMN, and 6 from FPN) and 60 edges that possessed a significant positive association with hallucinatory experience severity (p = 0.0192; **Figures 1B** and **1C**). That is, greater RSFC across edges was associated with greater hallucinatory experience severity. With the exception of a single edge between two CON parcels, the subnetwork is composed of inter-network connections, with 38 edges formed between CON and DMN parcels and 21 formed between CON and FPN parcels (**Figure 1C**). Major regions of involvement include the bilateral insula, ventromedial prefrontal cortex, inferior parietal lobe, and ventrolateral prefrontal cortex, as well as left anterior cingulate cortex and right hippocampal gyrus (see **Table S1**). Notably, the region with the highest degree centrality (at 14) is an FPN parcel of the left pre-supplementary motor area (pre-SMA). Follow-up regression analyses without GSR confirm that mean RSFC across edges is positively associated with hallucinatory experience severity (**Figure S2; Table S2**). Notably, the regression model predicting mean RSFC in DMN-CON edges calculated without GSR (**Figure S2A**) suggests that greater positive RSFC is associated with hallucinatory experience severity, rather than the possibility of a weaker anticorrelation between networks.

**Figure 1.**
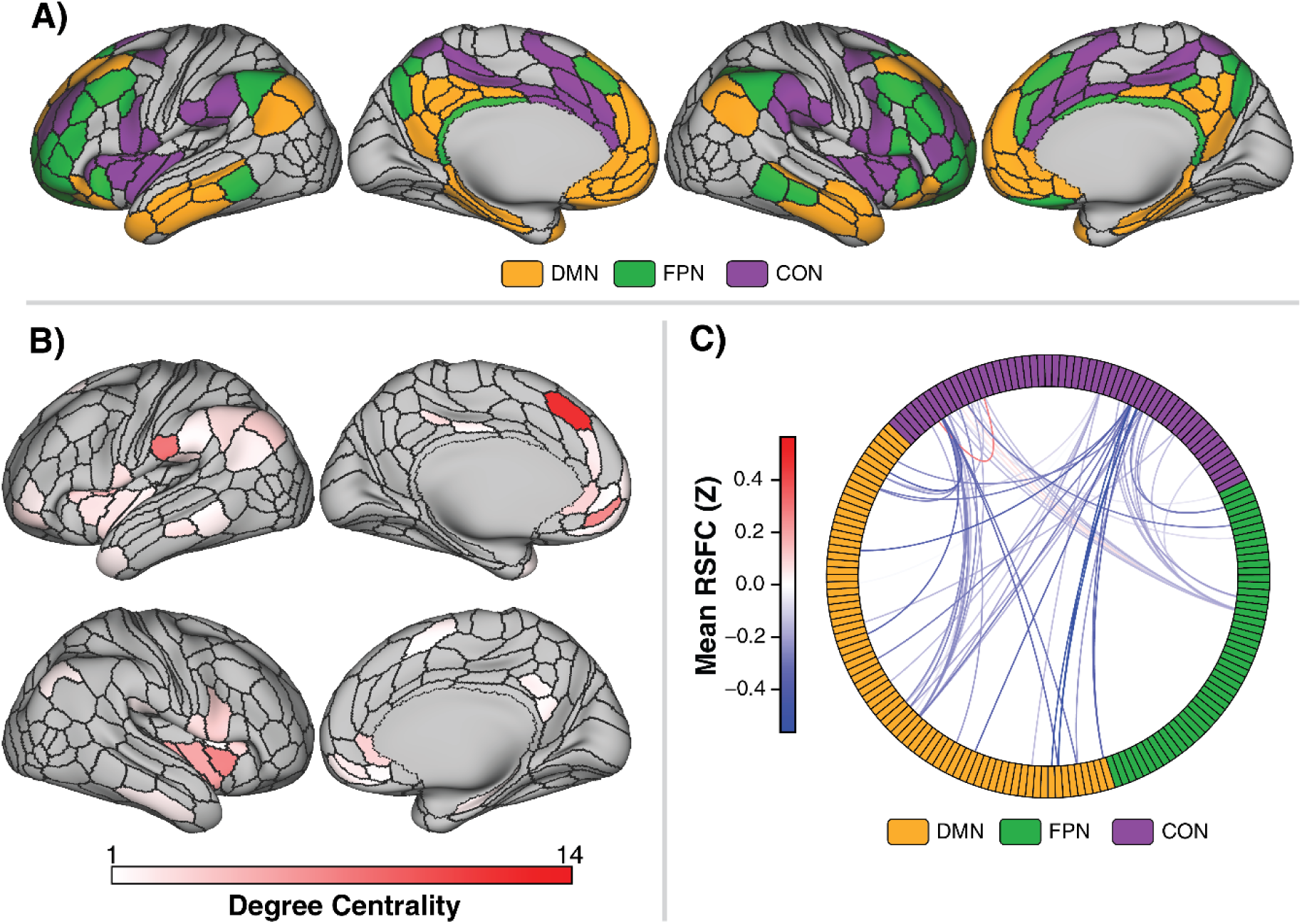
Connected Parcels Associated with Hallucination Severity. **A)** Regions of the Cole-Anticevic Brain Network Parcellation included in all connectivity analyses (183 cortical regions) from the Default Mode Network (DMN), Frontoparietal Network (FPN) and Cingulo-opercular Network (CON). **B)** Connected parcels found to be positively associated with Specific Psychotic Experiences Questionnaire (SPEQ) hallucination score via network-based statistic across 171 participants (p = 0.0192, family-wise error rate corrected), controlling for age, sex at birth, and stimulant medication status. Network-based statistic parameters included: 5,000 permutations, initial edge threshold of T = 3.1, and a cluster extent approach. Regions are displayed according to their degree centrality, which is the number of edges formed by each given region in the isolated subnetwork. **C)** Connectogram displaying all edges included in the isolated hallucination-specific subnetwork, colored according to the mean resting-state functional connectivity (RSFC) in each edge across participants.

Supplementary NBS analyses demonstrated that the association of increased RSFC with increased hallucinatory experience severity was robust to controlling for other self-reported SPEQ subscales (p = 0.008; **Figure S3, Table S3**) or total burden of psychopathology (p = 0.026; **Figure S4, Table S4**). Controlling for other SPEQ subscales, the resulting hallucination-associated subnetwork comprised 62 CON-DMN edges and 25 CON-FPN edges across 22 CON nodes, 34 DMN nodes, and 6 FPN nodes. Similarly, when controlling for psychopathology burden, the resulting subnetwork comprised 2 within-CON edges, 37 CON-DMN edges, and 13 CON-FPN edges across 17 CON nodes, 24 DMN nodes, and 3 FPN nodes.

### 3.3. Within-Network Connectivity and System Segregation

No within-network RSFC analysis showed a significant association with hallucination severity when corrected for multiple comparisons (**Figure 2A-C, Table S5**), although DMN within-network RSFC (across all DMN edges) was negatively associated with hallucination severity prior to correction for multiple comparisons (β* = -0.165, p = 0.030; **Figure 2A**). Poorer network-specific system segregation was significantly associated with worse hallucinatory experience severity in the DMN (β* = -0.143, p = 0.031; **Figure 2D, Table S6**) and CON (β* = - 0.218, p = 0.002; **Figure 2F, Table S6**). Further, global system segregation across all three networks was also negatively associated with hallucinatory experience severity (β* = -0.161, p = 0.015; **Figure 2G, Table S6**).

**Figure 2.**
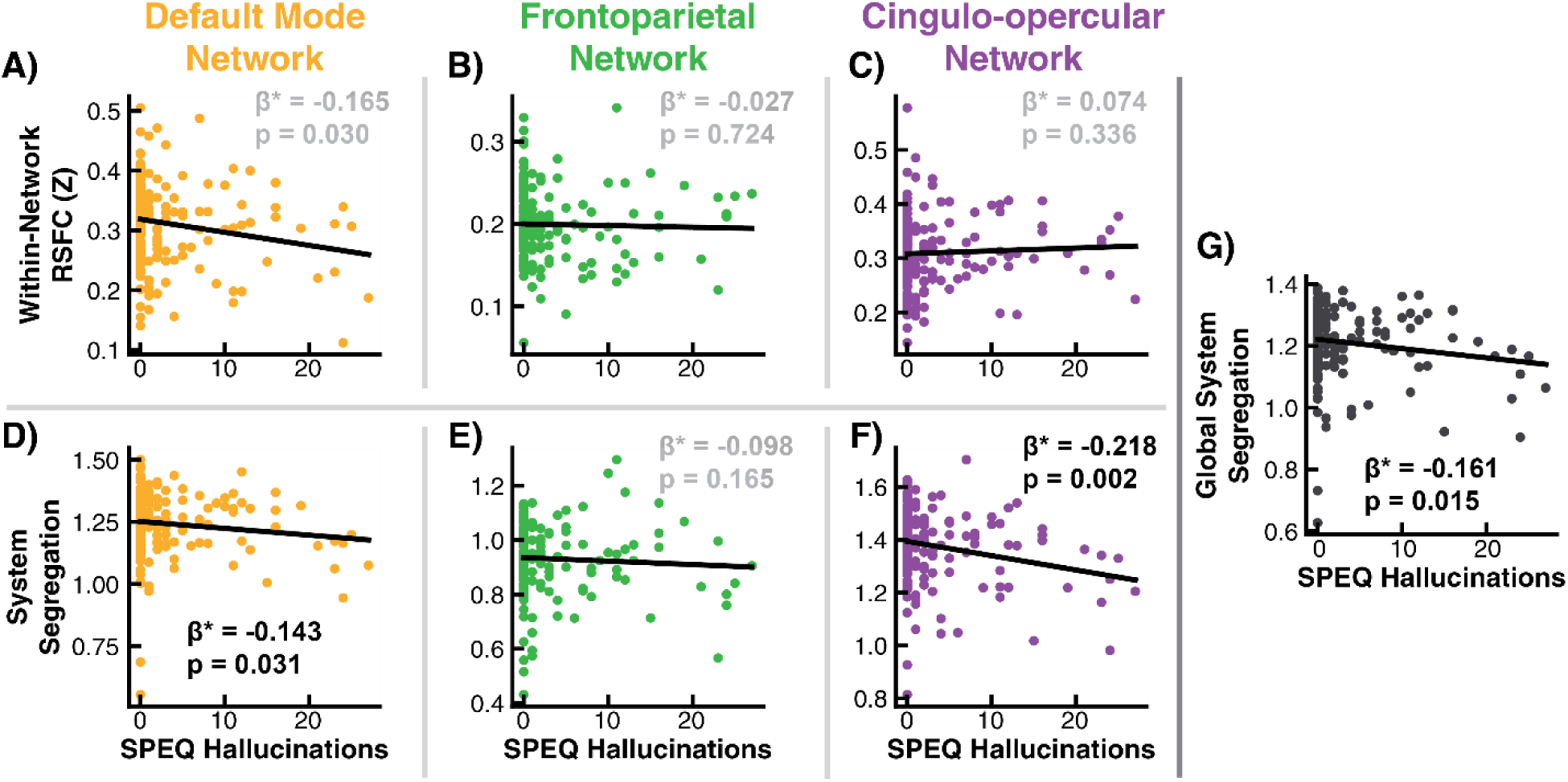
Associations between whole-network summary metrics and hallucination severity. A-C) Linear regressions predicting mean within-network resting-state functional connectivity (RSFC) across all edges within the Default Mode Network (A), Frontoparietal Network (B) and Cingulo-opercular Network (C) from Specific Psychotic Experiences Questionnaire (SPEQ) hallucination score, controlling for age, sex, and stimulant medication status. **D-G)** Linear regressions predicting network-specific system segregation for the Default Mode Network (D), Frontoparietal Network (E) and Cingulo-opercular Network (F), as well as global system segregation across all three networks (G) from Specific Psychotic Experiences Questionnaire (SPEQ) hallucination score, controlling for age, sex, and stimulant medication status. Standardized regression coefficients are denoted with β*. False discovery rate (FDR) correction was implemented separately for within-network RSFC models (A-C) and system segregation models (D-G). Standardized regression coefficients in black were significant following FDR correction.

### 3.4. NM-MRI Analyses

Mean VTA NM-MRI signal (see **Figure 3A** for ROI definition, **Figure 3B** for distribution of mean contrast values) was not associated with hallucination severity (**Table S7**). However, a significant negative association was observed between mean NM-MRI signal and the mean strength of RSFC in the subnetwork associated with hallucination severity (β* = -0.156, p = 0.038; **Figure 3C, Table S8**). An additional trend was observed towards a negative association between SNc NM-MRI signal and the hallucination-associated subnetwork (β* = -0.144, p = 0.067, **Table S9**).

**Figure 3.**
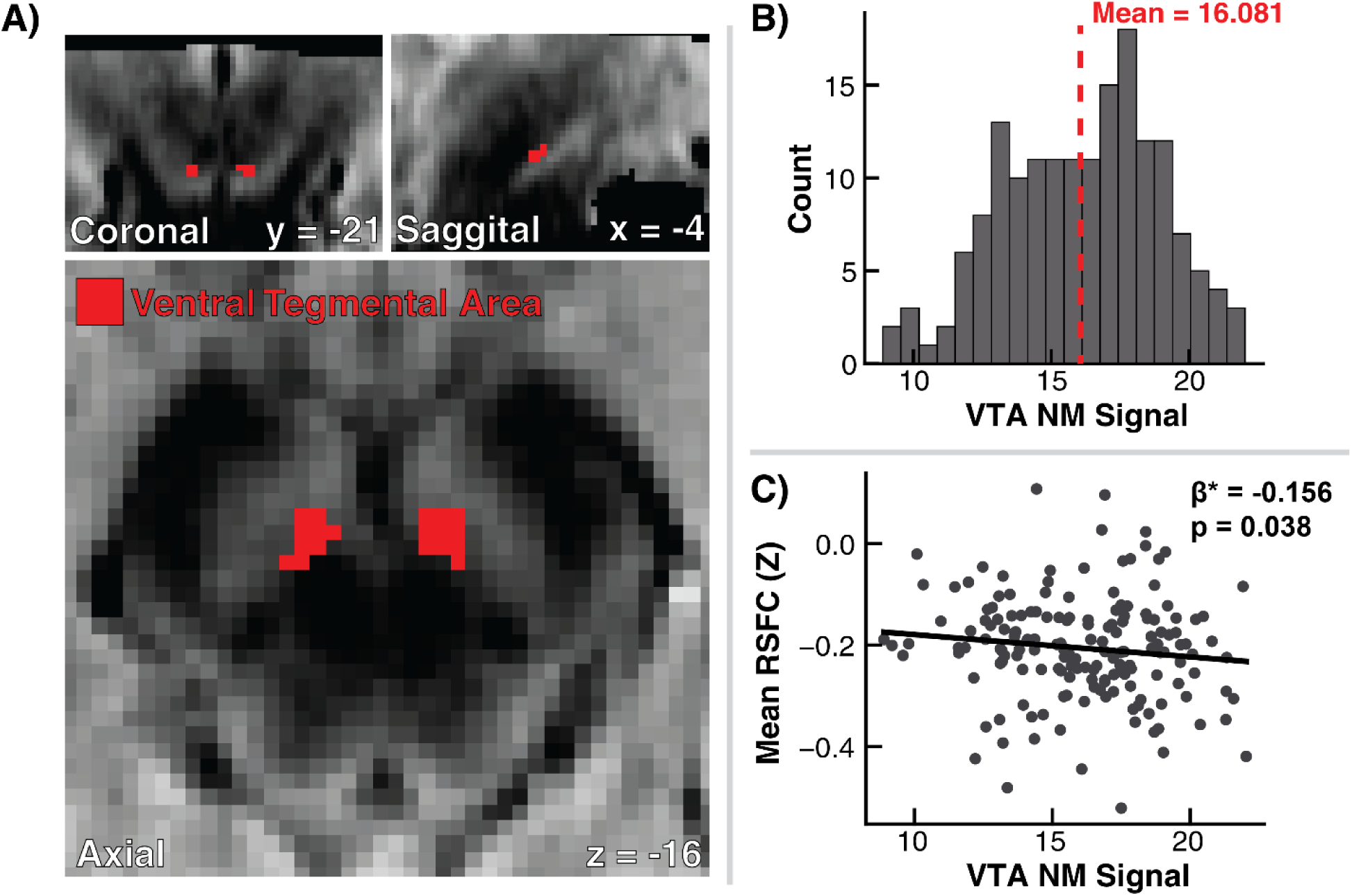
Neuromelanin Signal Associations with Hallucination Subnetwork Resting State Functional Connectivity and Hallucinatory Experience Severity. **A)** Ventral tegmental area (VTA) region of interest (29 voxels total; red overlay) used for all analyses of neuromelanin-sensitive magnetic resonance imaging (NM-MRI), overlaid on the group average NM signal image across all participants with NM-MRI data acquired (N = 165). **B)** Distribution of mean VTA NM signal values across all 165 participants, with a global mean value of 16.081 (red dashed line). **C)** Linear regression predicting mean resting-state functional connectivity (RSFC) across all edges in the hallucination-associated subnetwork from mean VTA NM signal, controlling for age, sex at birth, and stimulant medication status. Standardized regression coefficient is denoted with β*.

Follow-up network-specific analyses revealed a significant positive association between within-network DMN RSFC and VTA NM signal (β* = 0.224, p = 0.004, **Figure S5A, Table S10**). No additional significant associations were observed between VTA NM signal and within-network RSFC (**Figure S5B-C**) or system segregation (**Figure S5D-G, Table S11**).

### 3.5. Exploratory Network-Based Statistic Analysis

A follow-up exploratory NBS analysis aimed to clarify the spatial organization of VTA NM-MRI signal, which correlated with the hallucination-linked NBS network but not hallucination severity. This NBS identified a connected subnetwork of 29 parcels (1 CON, 1 FPN, and 27 DMN) and 36 edges in which greater RSFC was associated with greater VTA NM signal (**Figure 4, Table S12**). Of the 36 edges, 23 were between DMN parcels, 12 were between DMN parcels and a single CON region, and one was between a DMN parcel and FPN parcel. Major regions of involvement included the bilateral medial prefrontal cortex and posterior cingulate cortex/precuneus, with minimal involvement from bilateral lateral temporal lobe and left inferior parietal lobe. Intriguingly, this NM-associated subnetwork broadly corresponds to the “midline core” of the DMN^40^. In comparing this NM-associated subnetwork with the primary hallucination-associated subnetwork (shown in **Figure 1**), eight DMN parcels were found to be included in both networks (**Table S13**). However, there were zero overlapping RSFC edges between the two networks.

**Figure 4.**
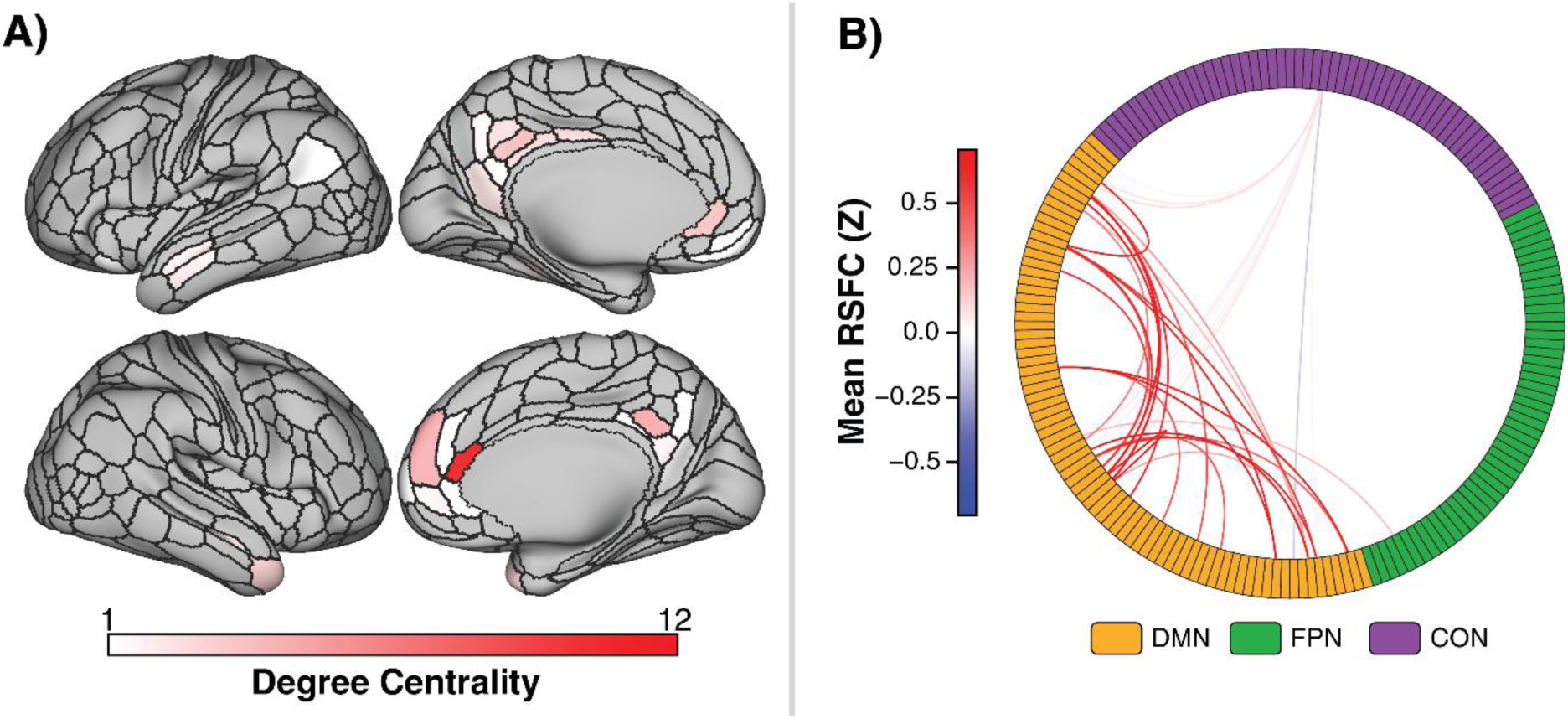
Connected Parcels Associated with Ventral Tegmental Area (VTA) Neuromelanin (NM) Signal. **A)** Connected parcels found to be positively associated with VTA NM signal via network-based statistic across 165 participants (p = 0.0492, family-wise error rate corrected), controlling for age, sex at birth, and stimulant medication status. Network-based statistic parameters included: 5,000 permutations, initial edge threshold of T = 3.1, and a cluster extent approach. Regions are displayed according to their degree centrality, which is the number of edges formed by each given region in the isolated subnetwork. **B)** Connectogram displaying all edges included in the isolated neuromelanin-specific subnetwork, colored according to the mean resting-state functional connectivity (RSFC) in each edge across participants.

## 4. Discussion

These findings provide evidence that subclinical hallucinatory experiences in late adolescence, a critical developmental period preceding the typical window for psychosis onset, are associated with altered cortical triple network organization. We demonstrated that hallucinatory experience severity was significantly associated with RSFC in a subnetwork of almost entirely CON-FPN and CON-DMN edges, consistent with reduced network segregation and greater coupling of FPN and DMN to CON. Similar topological patterns were observed when controlling for other PLE dimensions or total diagnostic burden, suggesting that this effect is not solely attributable to broad psychopathology burden or covariance with other PLEs. In contrast, although VTA NM-MRI signal, a measure of cumulative dopamine metabolism in the mesocortical dopaminergic projection, was negatively associated with mean RSFC in the hallucination-associated subnetwork, it was not directly associated with hallucinatory experience severity, indicating that dopamine-related midbrain signal may be linked more closely to large-scale network organization than to symptom severity directly. Exploratory NM-MRI beyond the VTA showed a similar but weaker association for SNc NM signal (p = 0.067), suggesting that the relationship between triple network RSFC and NM signal may extend across adjacent dopaminergic midbrain nuclei. Finally, follow-up analyses linked VTA NM-MRI signal specifically to RSFC in a largely spatially distinct subnetwork centered on DMN midline regions, particularly the midline core^40^. Together, these findings support a role for altered triple network organization in adolescent hallucinatory experiences, while suggesting that dopamine-related signal is associated with partially overlapping but topologically distinct patterns of cortical RSFC.

Broadly, these findings suggest partial continuity with, but not complete recapitulation of, adult models of psychosis. In schizophrenia, triple network dysfunction has been described in terms of both altered antagonism between DMN and FPN^41,42^ and aberrant salience network engagement, with prevalent models emphasizing dysfunction in an insula-/anterior cingulate-dominant salience system^11,43^ that incorrectly maps internal and external events and contributes to positive symptoms. In the present adolescent sample, hallucinatory experience severity was not associated with a direct reduction in DMN-FPN anticorrelation. Rather, it was associated with increased CON-DMN and CON-FPN coupling, together with reduced network segregation (**Figure 2D, F-G**). Thus, while the same large-scale networks relevant in adult psychosis appear to be implicated in adolescent hallucinatory experiences, we observe more prevalent connectivity alterations concerning CON regions that bridge networks, such as the anterior cingulate and insula, rather than a direct DMN-FPN abnormality. In addition, the left pre-SMA, a FPN region that is implicated broadly in cognitive effort, as well as in auditory imagery and inner speech generation^44^, was identified as a key network node. Further, pre-SMA has been specifically shown to be active during internal speech monitoring and error detection^45^ and has shown aberrant activity in persons experiencing both clinical and subclinical auditory hallucinations^46,47^. Greater connectivity of pre-SMA to CON regions responsible for salience attribution may therefore reflect an architectural vulnerability for source monitoring deficits, including the misattribution of internally generated stimuli.

These findings also have implications for understanding normal and abnormal neurodevelopment. The negative associations between hallucinatory experience severity and network segregation (**Figure 2D, F-G**) indicate that adolescents reporting more hallucinatory experiences show reduced differentiation among large-scale networks. Given that brain network segregation increases both functionally and structurally through childhood and adolescence^48,49^, one possible explanation is that adolescents who are prone to hallucinatory experiences show a pattern of large-scale network organization consistent with delayed or altered maturation. The NM-MRI findings should be interpreted more cautiously – although VTA NM-MRI signal was not directly associated with hallucinatory experience severity, it was related to mean connectivity in the hallucination-associated subnetwork and, more clearly, to within-DMN connectivity among midline core regions (**Figures 4A, 5**). Together, these findings suggest that dopamine-related midbrain signal may index a broader developmental process relevant to intrinsic network organization, whereas hallucinatory experiences may reflect one clinically meaningful correlate of reduced network segregation rather than a direct consequence of NM-MRI variation itself.

This interpretation is consistent with evidence that dopamine function facilitates the functional segregation and synaptic refinement of higher-order cognitive, motivational, and salience networks. There are several limitations of this work to acknowledge. First, while NBS is well-suited for making inferences about clusters of connected suprathreshold edges, it is not suitable for claiming that any singular edge in the identified cluster carries a statistically significant relationship with the independent variable. Accordingly, despite all edges in the hallucination-associated subnetwork individually passed an initial strict t-statistic threshold of 3.1, we cannot claim that any specific DMN-CON or FPN-CON edges disproportionately contribute to variations in hallucinatory experience severity. Second, as is typical with a community sample, our measure of hallucinatory experiences is positively skewed. That said, a large proportion of the sample reported at least some hallucinatory experiences (45.61%). Relatedly, the present hallucinatory phenotype should not be interpreted as equivalent in severity to hallucinations observed in schizophrenia, where hallucinations are often clinician-assessed rather than self-reported. This likely contributes to the modest effect sizes and partial, rather than total, corroboration of mechanistic accounts of hallucinations in schizophrenia reported throughout this study, notably the trend-level association between SNc NM-MRI signal and mean RSFC in the primary hallucination-associated subnetwork (**Figure 1, Table S9**).

A third limitation is that although we demonstrate that the relationship between altered triple network organization and hallucinatory experience severity is robust to controlling for total burden of psychopathology (**Figure S4**), this covariate is only a coarse index of diagnostic burden. Nevertheless, our community sample may also be viewed as a strength, ensuring generalizability of the effects while also replicating prior findings that hallucinatory experiences co-occur transdiagnostically with total burden of psychopathology^50^. Fourth, the cross-sectional nature of this work limits inferences about whether RSFC patterns drive the frequency and severity of hallucinatory experiences or simply passively correlate with them; longitudinal investigations are needed to test the temporal relationships between variables.

In summary, we extended prior evidence of relationships between DMN-CON-FPN RSFC integration to subclinical hallucinatory experiences in a previously unexamined age window preceding the age of psychosis onset. Further, we found that dopamine-related midbrain signal, indexed by VTA NM-MRI signal, was not associated with hallucinatory experience severity in adolescence, but with cortical connectivity patterns involving the hallucination-associated subnetwork and, more specifically, DMN regions corresponding to the midline core. The findings support a role for altered triple network organization in adolescents experiencing subclinical hallucinations, while suggesting that dopamine-related midbrain signal may be more closely linked to the broader development of triple network organization in this age range, rather than directly to symptom severity. Our work is a step toward identifying neural markers that may contribute to the development of high-risk psychosis phenotypes in late adolescence, which may ultimately help to inform future predictive models of psychosis conversion.

## 5. Data and Code Availability

Data used in this study is part of an ongoing longitudinal study that will be made available via the National Institute of Health Data Archive (NDA) upon its completion. All analysis code from this study can be made available upon request.

## Abbreviations

CABNP: Cole-Anticevic Brain Network Parcellation
CIFTI: Connectivity Informatics Technology Initiative
CON: Cingulo-opercular network
DMN: Default mode network
FPN: Frontoparietal network
GSR: Global signal regression
MP: Motion parameter
NBS: Network-based statistic
NM: neuromelanin
NM-MRI: Neuromelanin-sensitive magnetic resonance imaging
K-SADS: Kiddie Schedule for Affective Disorders and Schizophrenia
PET: Positron emission tomography
PLE: Psychotic-like experience
pre-SMA: Pre-supplementary motor area
ROI: Region of interest
RSFC: Resting-state functional connectivity
rs-fMRI: Resting-state functional magnetic resonance imaging
SN: Substantia nigra
SNc: Substantia nigra pars compacta
SPEQ: Specific Psychotic Experiences Questionnaire
VTA: Ventral tegmental area

