## Supplemental Material for "Distinct Connectivity Signatures of Hallucinatory Experiences and Neuromelanin Signal in Adolescents"

### Supplementary Methods

#### *Functional Connectivity Processing*

Following preprocessing, rs-fMRI data was mode-1000 normalized[1,2], linearly detrended, and mean-centered. Volumes acquired during periods of excess participant motion were removed using volume censoring with study-wide motion thresholds, measured by low-pass-filtered framewise displacement (LPF-FD), and run-wise thresholds for whole-brain signal fluctuation, using GEV-DV thresholding[3]. These fully data-driven thresholds were determined for task-based and resting-state data separately using the Multiband Censoring Optimization Tool (MCOT)[3]. Contiguous clusters of data shorter than 8 s in duration after censoring were removed; any runs with less than 1.5 minutes of remaining data were subsequently discarded. After censoring in either task, participants with fewer than 5 minutes of data total were excluded from further analyses. Further, we enforced that participants must have sufficient data in each of their two resting-state runs; our scan protocol was designed with alternating phase-encode directions, which have been shown to improve connectome reliability when averaged together over and above that of the anterior-posterior or poster-anterior directions alone[4].

Following censoring, runs were then filtered using a 0.009-0.08 Hz band-pass second-order zero-phase Butterworth filter, with censored time points replaced by linear interpolation prior to band-pass filtering before being discarded from analysis. The first and last 22 seconds of each timeseries were discarded to remove discontinuity artifacts present after filtering.

### Supplementary Figures

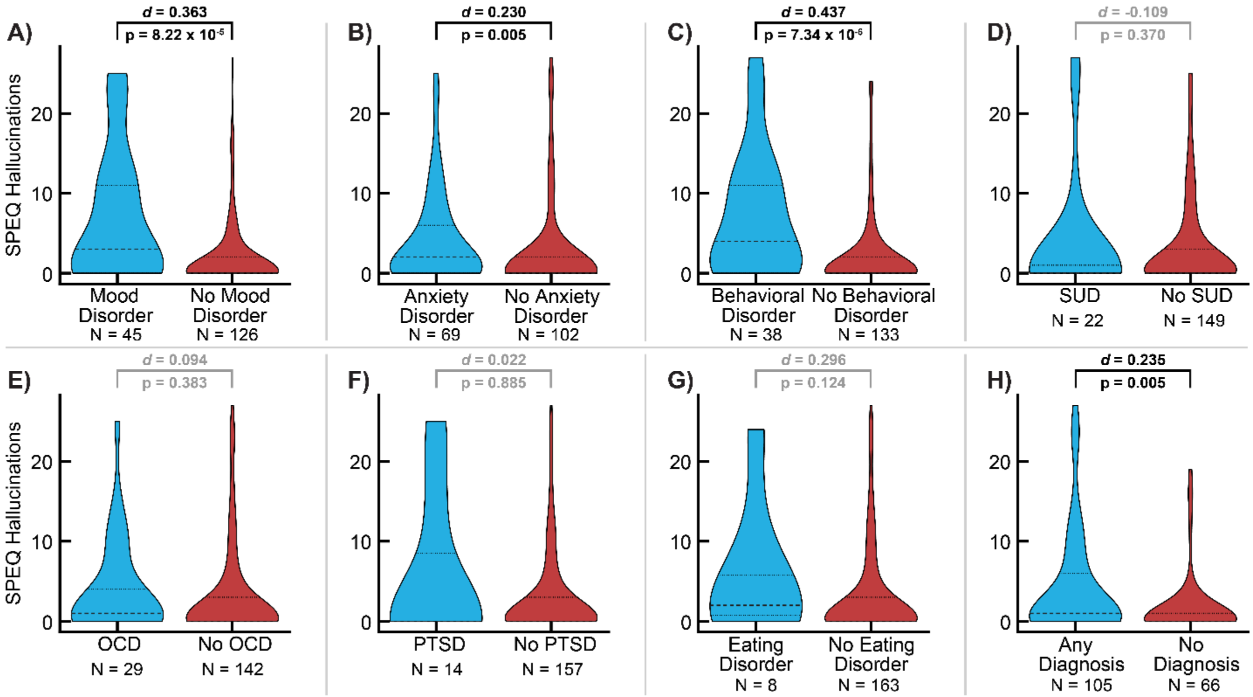


**Figure S1.** Comparisons of Specific Psychotic Experiences Questionnaire (SPEQ) Hallucination severity between diagnostic categories. Each panel displays separate distributions for a given diagnostic category (blue violin), as well as all participants without that diagnosis (red violin). Dashed lines within violins represent medians and quartiles. P-values were obtained via Mann-Whitney U-tests, and effect sizes were obtained via Cliff's delta (*d*). Black p-values were statistically significant following false discovery rate correction across all eight tests.

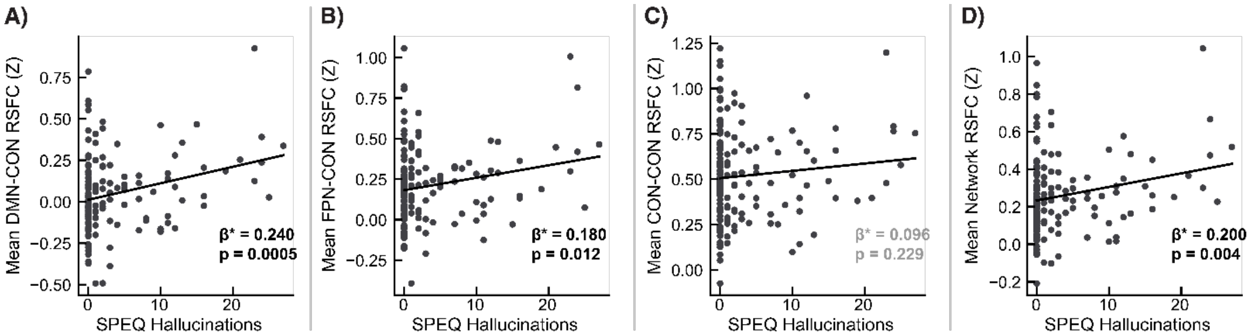


**Figure S2.** Associations between mean resting-state functional connectivity (RSFC) calculated without global signal regression in edges identified via Network Based Statistic that have a positive association with hallucination severity. Given that different edge types possess different biological baseline RSFC values without the use of global signal regression, mean RSFC was calculated separately for A) DMN-CON edges (38 edges total), B) FPN-CON edges (21 edges total), and 3) CON-CON edges (one edge). The association between mean RSFC across all 60 edges and hallucination severity is also shown in Panel D. Standardized regression coefficients are denoted with β*.

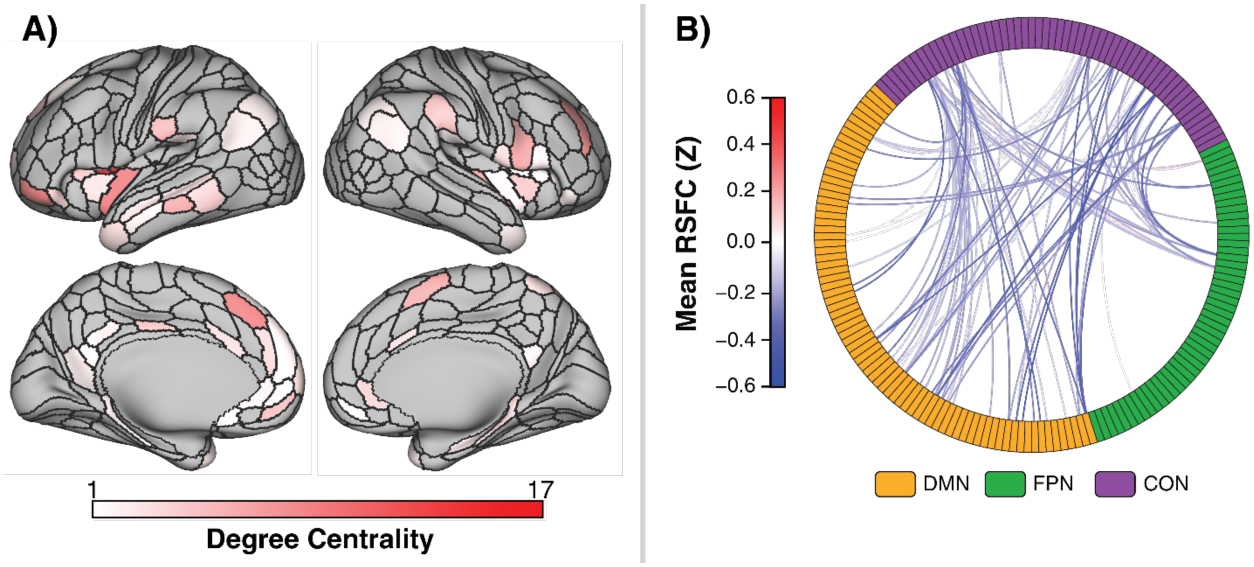


**Figure S3. A)** Connected parcels found to be positively associated with Specific Psychotic Experiences Questionnaire (SPEQ) hallucination score via network-based statistic across 171 participants (p = 0.008, family-wise error rate corrected), controlling for age, sex at birth, stimulant medication status, and SPEQ grandiosity, cognitive disorganization, anhedonia, and paranoia subscale scores. Network-based statistic parameters included: 5,000 permutations, initial edge threshold of T = 3.1, and a cluster extent approach. Regions are displayed according to their degree centrality, which is the number of edges formed by each given region in the isolated subnetwork. **B)** Connectogram displaying all edges included in the isolated subnetwork, colored according to the mean resting-state functional connectivity (RSFC) in each edge across participants.

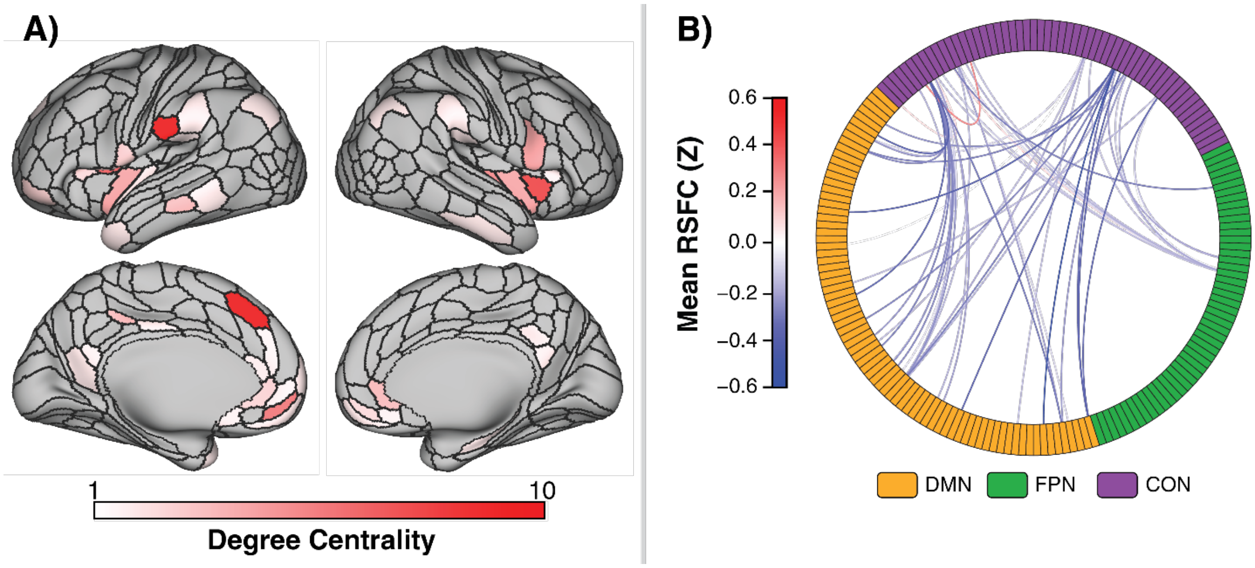


**Figure S4**. **A)** Connected parcels found to be positively associated with Specific Psychotic Experiences Questionnaire (SPEQ) hallucination score via network-based statistic across 171 participants (p = 0.026, family-wise error rate corrected), controlling for age, sex at birth, stimulant medication status, and total burden of psychopathology, measured as the number of diagnoses assigned via the Kiddie Schedule for Affective Disorders and Schizophrenia. Network-based statistic parameters included: 5,000 permutations, initial edge threshold of T = 3.1, and a cluster extent approach. Regions are displayed according to their degree centrality, which is the number of edges formed by each given region in the isolated subnetwork. **B)** Connectogram displaying all edges included in the isolated subnetwork, colored according to the mean resting-state functional connectivity (RSFC) in each edge across participants.

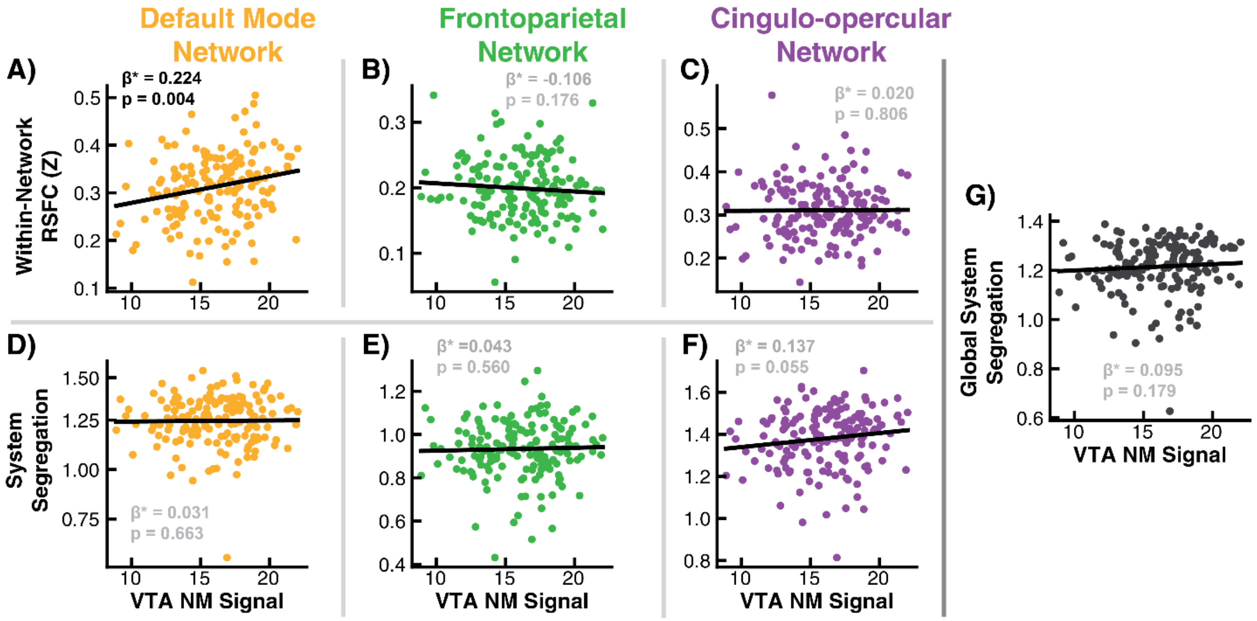


**Figure S5. Associations between whole-network summary metrics and neuromelanin (NM) signal. A-C)** Linear regressions predicting mean within-network resting-state functional connectivity (RSFC) across all edges within the Default Mode Network (A), Frontoparietal Network (B) and Cingulo-opercular Network (C) from mean NM signal in the ventral tegmental area (VTA), controlling for age, sex, and stimulant medication status. **D-G)** Linear regressions predicting network-specific system segregation for the Default Mode Network (D), Frontoparietal Network (E) and Cingulo-opercular Network (F), as well as global system segregation across all three networks (G) from VTA NM signal, controlling for age, sex, and stimulant medication status. Standardized regression coefficients are denoted with β*. False discovery rate (FDR) correction was implemented separately for within-network RSFC models (A-C) and system segregation models (D-G). Standardized regression coefficients in black were significant following FDR correction.

### Supplementary Tables

**Table S1.** Labels, anatomical regions, and degree centrality for each Cole-Anticevic Brain Network Parcellation (CAB-NP) parcel identified as a node in the hallucination-specific subnetwork identified via network-based statistic.

| CAB-NP Parcel Label | Anatomical Region | Degree Centrality |
|---|---|---|
| Cingulo-Opercular-06_R-Ctx | Supplementary/Cingulate Eye Field | 1 |
| Cingulo-Opercular-12_R-Ctx | Ventrolateral Prefrontal Cortex | 3 |
| Cingulo-Opercular-16_R-Ctx | Subcentral Area | 2 |
| Cingulo-Opercular-17_R-Ctx | Ventral Anterior Inferior Parietal Lobe | 1 |
| Cingulo-Opercular-18_R-Ctx | Posterior Insular Cortex | 6 |
| Cingulo-Opercular-19_R-Ctx | Frontal Operculum | 3 |
| Cingulo-Opercular-20_R-Ctx | Middle Insular Cortex | 8 |
| Cingulo-Opercular-21_R-Ctx | Frontal Operculum | 1 |
| Cingulo-Opercular-22_R-Ctx | Frontal Operculum | 1 |
| Cingulo-Opercular-25_R-Ctx | Posterior Insular Cortex | 6 |
| Cingulo-Opercular-43_L-Ctx | Subcentral Area | 2 |
| Cingulo-Opercular-44_L-Ctx | Ventral Anterior Inferior Parietal Lobe | 2 |
| Cingulo-Opercular-45_L-Ctx | Posterior Insular Cortex | 2 |
| Cingulo-Opercular-46_L-Ctx | Frontal Operculum | 3 |
| Cingulo-Opercular-47_L-Ctx | Middle Insular Cortex | 2 |
| Cingulo-Opercular-48_L-Ctx | Frontal Operculum | 2 |
| Cingulo-Opercular-49_L-Ctx | Frontal Operculum | 5 |
| Cingulo-Opercular-50_L-Ctx | Inferior Parietal Lobe | 9 |
| Cingulo-Opercular-51_L-Ctx | Inferior Parietal Lobe | 1 |
| Cingulo-Opercular-52_L-Ctx | Posterior Insular Cortex | 1 |
| Default-04_R-Ctx | Posterior Cingulate Gyrus | 1 |
| Default-06_R-Ctx | Posterior Cingulate Gyrus | 1 |
| Default-07_R-Ctx | Ventral Anterior Cingulate Gyrus | 3 |
| Default-09_R-Ctx | Ventromedial Prefrontal Cortex | 2 |
| Default-19_R-Ctx | Ventromedial Prefrontal Cortex | 1 |
| Default-24_R-Ctx | Hippocampal Gyrus | 1 |
| Default-30_R-Ctx | Inferior Temporal Gyrus | 1 |
| Default-32_R-Ctx | Inferior Parietal Lobe | 1 |
| Default-36_R-Ctx | Ventromedial Prefrontal Cortex | 1 |
| Default-40_L-Ctx | Posterior Cingulate Gyrus | 1 |
| Default-44_L-Ctx | Ventral Anterior Cingulate Gyrus | 3 |
| Default-45_L-Ctx | Dorsal Anterior Cingulate Gyrus | 2 |
| Default-46_L-Ctx | Ventromedial Prefrontal Cortex | 1 |
| Default-47_L-Ctx | Ventromedial Prefrontal Cortex | 8 |
| Default-54_L-Ctx | Medial Frontopolar Cortex | 1 |
| Default-57_L-Ctx | Ventromedial Prefrontal Cortex | 1 |
| Default-66_L-Ctx | Temporal Pole | 1 |
| Default-69_L-Ctx | Inferior Parietal Lobe | 1 |
| Default-70_L-Ctx | Inferior Parietal Lobe | 3 |
| Default-73_L-Ctx | Posterior Cingulate Gyrus | 2 |
| Default-77_L-Ctx | Middle Temporal Gyrus | 2 |
| Frontoparietal-32_L-Ctx | Pre-Supplementary Motor Area | 14 |
| Frontoparietal-34_L-Ctx | Lateral Frontopolar Cortex | 2 |
| Frontoparietal-43_L-Ctx | Supplementary Eye Field | 1 |
| Frontoparietal-45_L-Ctx | Middle Temporal Gyrus | 1 |
| Frontoparietal-48_L-Ctx | Supramarginal Gyrus | 2 |
| Frontoparietal-50_L-Ctx | Ventrolateral Prefrontal Cortex | 1 |

**Table S2.** Regression models predicting mean resting-state functional connectivity (RSFC) calculated without global signal regression in edges identified via Network Based Statistic from the following predictors: intercept, age, sex at birth, stimulant medication status, and Specific Psychotic Experiences Questionnaire Hallucination score.

| **Y = Default Mode Network-Cingulo-Opercular Network RSFC[a] (38 edges)** | | | | | |
|---|---|---|---|---|---|
| **Effect** | **Estimate (*B*)** | ***SE* (*B*)** | **β*** | **$t_{159}$** | **p** |
| Intercept | 0.033 | 0.015 | - | -0.816 | 0.416 |
| Age | -0.036 | 0.015 | -0.165 | -2.369 | 0.019 |
| Sex at Birth (Female) | -0.033 | 0.015 | -0.148 | -2.184 | 0.030 |
| Stimulant Medication | -0.006 | 0.015 | 0.028 | 0.405 | 0.686 |
| SPEQ Hallucinations | 0.054 | 0.015 | 0.240 | 3.542 | 0.0005 |
| **Y = Frontoparietal Network-Cingulo-Opercular Network RSFC[b] (21 edges)** | | | | | |
| **Effect** | **Estimate (*B*)** | ***SE* (*B*)** | **β*** | **$t_{159}$** | **p** |
| Intercept | 0.194 | 0.016 | - | -0.850 | 0.397 |
| Age | -0.036 | 0.016 | -0.155 | -2.197 | 0.030 |
| Sex at Birth (Female) | -0.023 | 0.016 | -0.099 | -1.401 | 0.163 |
| Stimulant Medication | 0.021 | 0.016 | 0.092 | 1.297 | 0.197 |
| SPEQ Hallucinations | 0.041 | 0.016 | 0.180 | 2.555 | 0.012 |
| **Y = Cingulo-Opercular Network Within-Network RSFC[c] (1 edge)** | | | | | |
| **Effect** | **Estimate (*B*)** | ***SE* (*B*)** | **β*** | **$t_{159}$** | **p** |
| Intercept | 0.508 | 0.020 | - | -0.566 | 0.572 |
| Age | -0.016 | 0.020 | -0.065 | -0.822 | 0.412 |
| Sex at Birth (Female) | 0.033 | 0.020 | 0.134 | 1.689 | 0.093 |
| Stimulant Medication | -0.013 | 0.020 | -0.051 | -0.636 | 0.526 |
| SPEQ Hallucinations | 0.024 | 0.020 | 0.096 | 1.208 | 0.229 |
| **Y = Mean RSFC across all NBS-identified edges[d] (60 edges)** | | | | | |
| **Effect** | **Estimate (*B*)** | ***SE* (*B*)** | **β*** | **$t_{159}$** | **p** |
| Intercept | 0.240 | 0.014 | - | -1.209 | 0.229 |
| Age | -0.029 | 0.014 | -0.142 | -2.066 | 0.041 |
| Sex at Birth (Female) | -0.008 | 0.014 | -0.038 | -0.544 | 0.587 |
| Stimulant Medication | 0.007 | 0.014 | 0.034 | 0.489 | 0.626 |
| SPEQ Hallucinations | 0.040 | 0.014 | 0.200 | 2.910 | 0.004 |

[a]N = 159, Error degrees of freedom = 154, R-squared = 0.132, Adjusted R-squared = 0.110, F-statistic versus constant model = 5.87, p-value = 0.0002.
[b]N = 159, Error degrees of freedom = 154, R-squared = 0.090, Adjusted R-squared = 0.066, F-statistic versus constant model = 3.80, p-value = 0.006.
[c]N = 159, Error degrees of freedom = 154, R-squared = 0.040, Adjusted R-squared = 0.015, F-statistic versus constant model = 1.60, p-value = 0.177.
[d]N = 159, Error degrees of freedom = 154, R-squared = 0.087, Adjusted R-squared = 0.064, F-statistic versus constant model = 3.69, p-value = 0.007.
Model coefficients were estimated using robust regression with Huber weighting.
*Abbreviations*: RSFC, resting-state functional connectivity; SE, standard error; β*, standardized regression coefficient; SPEQ, Specific Psychotic Experiences Questionnaire; NBS, Network-based statistic.

**Table S3.** Labels, anatomical regions, and degree centrality for each Cole-Anticevic Brain Network Parcellation (CAB-NP) parcel identified as a node in the hallucination-specific subnetwork identified via network-based statistic controlling for other self-reported psychotic-like experience scales from the Specific Psychotic Experiences Questionnaire.

| CAB-NP Parcel Label | Anatomical Region | Degree Centrality |
|---|---|---|
| Cingulo-Opercular-06_R-Ctx | Supplementary/Cingulate Eye Field | 6 |
| Cingulo-Opercular-08_R-Ctx | Precuneus | 2 |
| Cingulo-Opercular-10_R-Ctx | Dorsal Anterior Cingulate | 2 |
| Cingulo-Opercular-12_R-Ctx | Ventrolateral Prefrontal Cortex | 6 |
| Cingulo-Opercular-14_R-Ctx | Dorsolateral Prefrontal Cortex | 8 |
| Cingulo-Opercular-16_R-Ctx | Subcentral Area | 2 |
| Cingulo-Opercular-17_R-Ctx | Ventral Anterior Inferior Parietal Lobe | 1 |
| Cingulo-Opercular-18_R-Ctx | Posterior Insular Cortex | 1 |
| Cingulo-Opercular-19_R-Ctx | Frontal Operculum | 1 |
| Cingulo-Opercular-20_R-Ctx | Middle Insular Cortex | 4 |
| Cingulo-Opercular-21_R-Ctx | Frontal Operculum | 1 |
| Cingulo-Opercular-22_R-Ctx | Frontal Operculum | 1 |
| Cingulo-Opercular-24_R-Ctx | Inferior Parietal Lobe | 4 |
| Cingulo-Opercular-25_R-Ctx | Posterior Insular Cortex | 6 |
| Cingulo-Opercular-26_R-Ctx | Frontal Operculum | 1 |
| Cingulo-Opercular-38_L-Ctx | Dorsal Anterior Cingulate | 2 |
| Cingulo-Opercular-44_L-Ctx | Ventral Anterior Inferior Parietal Lobe | 2 |
| Cingulo-Opercular-45_L-Ctx | Posterior Insular Cortex | 9 |
| Cingulo-Opercular-46_L-Ctx | Frontal Operculum | 4 |
| Cingulo-Opercular-47_L-Ctx | Middle Insular Cortex | 2 |
| Cingulo-Opercular-49_L-Ctx | Frontal Operculum | 17 |
| Cingulo-Opercular-50_L-Ctx | Inferior Parietal Lobe | 5 |
| Default-04_R-Ctx | Posterior Cingulate Gyrus | 1 |
| Default-07_R-Ctx | Ventral Anterior Cingulate Gyrus | 3 |
| Default-09_R-Ctx | Ventromedial Prefrontal Cortex | 1 |
| Default-15_R-Ctx | Superior Frontal Gyrus | 1 |
| Default-17_R-Ctx | Ventrolateral Prefrontal Cortex | 1 |
| Default-18_R-Ctx | Superior Frontal Gyrus | 1 |
| Default-23_R-Ctx | Hippocampal Gyrus | 2 |
| Default-24_R-Ctx | Hippocampal Gyrus | 1 |
| Default-28_R-Ctx | Temporal Pole | 1 |
| Default-31_R-Ctx | Inferior Parietal Lobe | 1 |
| Default-32_R-Ctx | Inferior Parietal Lobe | 1 |
| Default-39_L-Ctx | Parieto-occipital Sulcus | 1 |
| Default-40_L-Ctx | Posterior Cingulate Gyrus | 4 |
| Default-41_L-Ctx | Retrosplenial Cortex | 1 |
| Default-43_L-Ctx | Posterior Cingulate Gyrus | 1 |
| Default-44_L-Ctx | Ventral Anterior Cingulate Gyrus | 1 |
| Default-45_L-Ctx | Dorsal Anterior Cingulate Gyrus | 3 |
| Default-46_L-Ctx | Ventromedial Prefrontal Cortex | 1 |
| Default-47_L-Ctx | Ventromedial Prefrontal Cortex | 5 |
| Default-50_L-Ctx | Superior Frontal Gyrus | 1 |
| Default-51_L-Ctx | Dorsomedial Prefrontal Cortex | 1 |
| Default-52_L-Ctx | Superior Frontal Gyrus | 1 |
| Default-53_L-Ctx | Superior Frontal Gyrus | 4 |
| Default-54_L-Ctx | Medial Frontopolar Cortex | 2 |
| Default-55_L-Ctx | Ventrolateral Prefrontal Cortex | 3 |
| Default-56_L-Ctx | Superior Frontal Gyrus | 1 |
| Default-62_L-Ctx | Hippocampal Gyrus | 1 |

| | | |
|---|---|---|
| Default-65_L-Ctx | Superior Temporal Sulcus | 2 |
| Default-66_L-Ctx | Temporal Pole | 2 |
| Default-67_L-Ctx | Superior Temporal Sulcus | 1 |
| Default-69_L-Ctx | Inferior Parietal Lobe | 1 |
| Default-70_L-Ctx | Inferior Parietal Lobe | 1 |
| Default-71_L-Ctx | Inferior Temporal Sulcus | 1 |
| Default-74_L-Ctx | Ventral Anterior Cingulate Gyrus | 1 |
| Default-76_L-Ctx | Superior Temporal Sulcus | 2 |
| Default-77_L-Ctx | Middle Temporal Gyrus | 6 |
| Frontoparietal-08_R-Ctx | Ventrolateral Prefrontal Cortex | 1 |
| Frontoparietal-32_L-Ctx | Pre-Supplementary Motor Area | 9 |
| Frontoparietal-34_L-Ctx | Lateral Frontopolar Cortex | 9 |
| Frontoparietal-39_L-Ctx | Lateral Frontopolar Cortex | 1 |
| Frontoparietal-45_L-Ctx | Middle Temporal Gyrus | 3 |
| Frontoparietal-49_L-Ctx | Lateral Frontopolar Cortex | 2 |

**Table S4.** Labels, anatomical regions, and degree centrality for each Cole-Anticevic Brain Network Parcellation (CAB-NP) parcel identified as a node in the hallucination-specific subnetwork identified via network-based statistic controlling for total burden of psychopathology, measured as the total number of diagnoses recorded from the Computerized Kiddie Schedule for Affective Disorders and Schizophrenia.

| CAB-NP Parcel Label | Anatomical Region | Degree Centrality |
|---|---|---|
| Cingulo-Opercular-12_R-Ctx | Ventrolateral Prefrontal Cortex | 4 |
| Cingulo-Opercular-17_R-Ctx | Ventral Anterior Inferior Parietal Lobe | 1 |
| Cingulo-Opercular-18_R-Ctx | Posterior Insular Cortex | 4 |
| Cingulo-Opercular-19_R-Ctx | Frontal Operculum | 1 |
| Cingulo-Opercular-20_R-Ctx | Middle Insular Cortex | 8 |
| Cingulo-Opercular-24_R-Ctx | Inferior Parietal Lobe | 1 |
| Cingulo-Opercular-25_R-Ctx | Posterior Insular Cortex | 5 |
| Cingulo-Opercular-43_L-Ctx | Subcentral Area | 2 |
| Cingulo-Opercular-44_L-Ctx | Ventral Anterior Inferior Parietal Lobe | 1 |
| Cingulo-Opercular-45_L-Ctx | Posterior Insular Cortex | 4 |
| Cingulo-Opercular-46_L-Ctx | Frontal Operculum | 2 |
| Cingulo-Opercular-48_L-Ctx | Frontal Operculum | 1 |
| Cingulo-Opercular-49_L-Ctx | Frontal Operculum | 7 |
| Cingulo-Opercular-50_L-Ctx | Inferior Parietal Lobe | 10 |
| Cingulo-Opercular-51_L-Ctx | Inferior Parietal Lobe | 1 |
| Cingulo-Opercular-52_L-Ctx | Posterior Insular Cortex | 1 |
| Cingulo-Opercular-56_L-Ctx | Dorsal Anterior Cingulate | 1 |
| Default-04_R-Ctx | Posterior Cingulate Gyrus | 1 |
| Default-06_R-Ctx | Posterior Cingulate Gyrus | 1 |
| Default-07_R-Ctx | Ventral Anterior Cingulate Gyrus | 3 |
| Default-09_R-Ctx | Ventromedial Prefrontal Cortex | 2 |
| Default-19_R-Ctx | Ventromedial Prefrontal Cortex | 1 |
| Default-24_R-Ctx | Hippocampal Gyrus | 1 |
| Default-30_R-Ctx | Inferior Temporal Gyrus | 1 |
| Default-32_R-Ctx | Inferior Parietal Lobe | 1 |
| Default-36_R-Ctx | Ventromedial Prefrontal Cortex | 1 |
| Default-39_L-Ctx | Parieto-occipital Sulcus | 1 |
| Default-40_L-Ctx | Posterior Cingulate Gyrus | 1 |
| Default-41_L-Ctx | Retrosplenial Cortex | 1 |
| Default-44_L-Ctx | Ventral Anterior Cingulate Gyrus | 2 |
| Default-45_L-Ctx | Dorsal Anterior Cingulate Gyrus | 1 |
| Default-46_L-Ctx | Ventromedial Prefrontal Cortex | 1 |
| Default-47_L-Ctx | Ventromedial Prefrontal Cortex | 6 |
| Default-53_L-Ctx | Superior Frontal Gyrus | 1 |
| Default-54_L-Ctx | Medial Frontopolar Cortex | 1 |
| Default-57_L-Ctx | Ventromedial Prefrontal Cortex | 1 |
| Default-66_L-Ctx | Temporal Pole | 1 |
| Default-70_L-Ctx | Inferior Parietal Lobe | 1 |
| Default-73_L-Ctx | Posterior Cingulate Gyrus | 3 |
| Default-74_L-Ctx | Ventral Anterior Cingulate Gyrus | 1 |
| Default-77_L-Ctx | Middle Temporal Gyrus | 3 |
| Frontoparietal-32_L-Ctx | Pre-Supplementary Motor Area | 10 |
| Frontoparietal-34_L-Ctx | Lateral Frontopolar Cortex | 2 |
| Frontoparietal-45_L-Ctx | Middle Temporal Gyrus | 1 |

**Table S5.** Regression models predicting within-network resting-state functional connectivity in each of the default mode network, frontoparietal network, and cingulo-opercular network from the following predictors: intercept, age, sex at birth, stimulant medication status, and Specific Psychotic Experiences Questionnaire Hallucination score.

| Y = Default Mode Network Within-Network RSFC[a] | | | | | |
|---|---|---|---|---|---|
| **Effect** | **Estimate (*B*)** | ***SE* (*B*)** | **β*** | **$t_{171}$** | **p** |
| Intercept | 0.310 | 0.005 | - | 0.081 | 0.936 |
| Age | 0.004 | 0.005 | 0.060 | 0.794 | 0.428 |
| Sex at Birth (Female) | 0.012 | 0.005 | 0.017 | 2.267 | 0.025 |
| Stimulant Medication | -0.001 | 0.005 | -0.008 | -0.112 | 0.911 |
| SPEQ Hallucinations | -0.012 | 0.005 | -0.165 | -2.188 | 0.030 |
| **Y = Frontoparietal Network Within-Network RSFC[b]** | | | | | |
| **Effect** | **Estimate (*B*)** | ***SE* (*B*)** | **β*** | **$t_{171}$** | **p** |
| Intercept | 0.199 | 0.003 | - | -0.187 | 0.851 |
| Age | -0.004 | 0.003 | -0.096 | -1.276 | 0.204 |
| Sex at Birth (Female) | -0.007 | 0.003 | 0.157 | 2.093 | 0.038 |
| Stimulant Medication | -0.002 | 0.003 | -0.044 | -0.582 | 0.562 |
| SPEQ Hallucinations | -0.001 | 0.003 | -0.027 | -0.354 | 0.724 |
| **Y = Cingulo-opercular Network Within-Network RSFC[c]** | | | | | |
| **Effect** | **Estimate (*B*)** | ***SE* (*B*)** | **β*** | **$t_{171}$** | **p** |
| Intercept | 0.309 | 0.005 | - | -0.236 | 0.814 |
| Age | 0.001 | 0.005 | 0.019 | 0.242 | 0.809 |
| Sex at Birth (Female) | -0.001 | 0.005 | -0.023 | -0.305 | 0.761 |
| Stimulant Medication | -0.001 | 0.005 | -0.011 | -0.146 | 0.884 |
| SPEQ Hallucinations | 0.004 | 0.005 | 0.074 | 0.965 | 0.336 |

[a]N = 171, Error degrees of freedom = 166, R-squared = 0.059, Adjusted R-squared = 0.037, F-statistic versus constant model = 2.62, p-value = 0.037.

[b]N = 171, Error degrees of freedom = 166, R-squared = 0.041, Adjusted R-squared = 0.018, F-statistic versus constant model = 1.79, p-value = 0.134.

[c]N = 171, Error degrees of freedom = 166, R-squared = 0.007, Adjusted R-squared = -0.017, F-statistic versus constant model = 0.297, p-value = 0.880.

Model coefficients were estimated using robust regression with Huber weighting.

*Abbreviations*: RSFC, resting-state functional connectivity; SE, standard error; β*, standardized regression coefficient; SPEQ, Specific Psychotic Experiences Questionnaire.

**Table S6.** Regression models predicting network-specific system segregation in each of the default mode network, frontoparietal network, and cingulo-opercular network, as well as global system segregation, from the following predictors: intercept, age, sex at birth, stimulant medication status, and Specific Psychotic Experiences Questionnaire Hallucination score.

| **Y = Default Mode Network-specific System Segregation[a]** | | | | | |
|---|---|---|---|---|---|
| **Effect** | **Estimate (*B*)** | ***SE* (*B*)** | **β*** | **$t_{171}$** | **p** |
| Intercept | 1.251 | 0.008 | - | 1.058 | 0.292 |
| Age | 0.006 | 0.008 | 0.045 | 0.689 | 0.492 |
| Sex at Birth (Female) | -0.022 | 0.008 | -0.174 | -2.636 | 0.006 |
| Stimulant Medication | -0.006 | 0.008 | -0.047 | -0.708 | 0.480 |
| SPEQ Hallucinations | -0.018 | 0.008 | -0.143 | -2.177 | 0.031 |
| **Y = Frontoparietal Network-specific System Segregation[b]** | | | | | |
| **Effect** | **Estimate (*B*)** | ***SE* (*B*)** | **β*** | **$t_{171}$** | **p** |
| Intercept | 0.939 | 0.009 | - | 0.838 | 0.403 |
| Age | 0.005 | 0.009 | 0.038 | 0.534 | 0.594 |
| Sex at Birth (Female) | -0.012 | 0.009 | -0.087 | -1.238 | 0.217 |
| Stimulant Medication | -0.009 | 0.009 | -0.065 | -0.916 | 0.361 |
| SPEQ Hallucinations | -0.013 | 0.009 | -0.098 | -1.396 | 0.165 |
| **Y = Cingulo-opercular Network-specific System Segregation[c]** | | | | | |
| **Effect** | **Estimate (*B*)** | ***SE* (*B*)** | **β*** | **$t_{171}$** | **p** |
| Intercept | 1.386 | 0.010 | - | 0.985 | 0.326 |
| Age | 0.016 | 0.010 | 0.117 | 1.700 | 0.091 |
| Sex at Birth (Female) | 0.033 | 0.010 | 0.237 | 3.438 | 0.0007 |
| Stimulant Medication | -0.004 | 0.010 | -0.029 | -0.415 | 0.679 |
| SPEQ Hallucinations | -0.031 | 0.010 | -0.217 | -3.170 | 0.002 |
| **Y = Global System Segregation[d]** | | | | | |
| **Effect** | **Estimate (*B*)** | ***SE* (*B*)** | **β*** | **$t_{171}$** | **p** |
| Intercept | 1.224 | 0.007 | - | 1.708 | 0.089 |
| Age | 0.009 | 0.007 | 0.075 | 1.146 | 0.253 |
| Sex at Birth (Female) | -0.003 | 0.007 | -0.024 | -0.365 | 0.715 |
| Stimulant Medication | -0.005 | 0.007 | -0.045 | -0.693 | 0.489 |
| SPEQ Hallucinations | -0.018 | 0.007 | -0.160 | -2.454 | 0.015 |

[a]N = 171, Error degrees of freedom = 166, R-squared = 0.076, Adjusted R-squared = 0.054, F-statistic versus constant model = 3.43, p-value = 0.010.

[b]N = 171, Error degrees of freedom = 166, R-squared = 0.030, Adjusted R-squared = 0.007, F-statistic versus constant model = 1.28, p-value = 0.280.

[c]N = 171, Error degrees of freedom = 166, R-squared = 0.133, Adjusted R-squared = 0.112, F-statistic versus constant model = 6.36, p-value = $8.66 \times 10^{-5}$.

[d]N = 171, Error degrees of freedom = 166, R-squared = 0.061, Adjusted R-squared = 0.038, F-statistic versus constant model = 2.70, p-value = 0.033.

Model coefficients were estimated using robust regression with Huber weighting.

*Abbreviations*: SE, standard error; β*, standardized regression coefficient; SPEQ, Specific Psychotic Experiences Questionnaire.

**Table S7.** Regression model predicting Specific Psychotic Experiences Questionnaire Hallucination score using the following predictors: intercept, age, sex at birth, stimulant medication status, and mean neuromelanin signal in the ventral tegmental area.

| **Effect** | **Estimate (*B*)** | ***SE* (*B*)** | **β*** | **$t_{165}$** | **p** |
|---|---|---|---|---|---|
| Intercept | 1.411 | 0.236 | -0.301 | -7.340 | $1.020 \times 10^{-11}$ |
| Age | 0.035 | 0.239 | 0.006 | 0.145 | 0.885 |
| Sex at Birth (Female) | -0.076 | 0.241 | -0.013 | -0.317 | 0.752 |
| Stimulant Medication | -0.126 | 0.238 | -0.022 | -0.529 | 0.597 |
| VTA NM Signal | 0.044 | 0.241 | -0.008 | 0.183 | 0.855 |

*Note.* N = 165, Error degrees of freedom = 160, R-squared = 0.253, Adjusted R-squared = 0.235, F-statistic versus constant model = 13.6, p-value = $1.51 \times 10^{-9}$.

Model coefficients were estimated using robust regression with Huber weighting.

*Abbreviations*: SE, standard error; β*, standardized regression coefficient; VTA, ventral tegmental area; NM, neuromelanin.

**Table S8.** Regression model predicting mean resting-state functional connectivity across edges identified to be associated with hallucination severity via network-based statistic using the following predictors: intercept, age, sex at birth, stimulant medication status, and mean neuromelanin signal in the ventral tegmental area.

| **Effect** | **Estimate (*B*)** | ***SE* (*B*)** | **β*** | **$t_{165}$** | **p** |
|---|---|---|---|---|---|
| Intercept | -0.206 | 0.008 | - | 0.006 | 0.995 |
| Age | -0.007 | 0.008 | -0.071 | -0.962 | 0.338 |
| Sex at Birth (Female) | -0.006 | 0.008 | -0.059 | -0.795 | 0.428 |
| Stimulant Medication | 0.0003 | 0.008 | 0.003 | 0.039 | 0.969 |
| VTA NM Signal | -0.016 | 0.008 | -0.156 | -2.088 | 0.038 |

*Note.* N = 165, Error degrees of freedom = 160, R-squared = 0.036, Adjusted R-squared = 0.012, F-statistic versus constant model = 1.50, p-value = 0.204.
Model coefficients were estimated using robust regression with Huber weighting.
*Abbreviations*: SE, standard error; β*, standardized regression coefficient; VTA, ventral tegmental area; NM, neuromelanin.

**Table S9.** Regression model predicting mean resting-state functional connectivity across edges identified to be associated with hallucination severity via network-based statistic using the following predictors: intercept, age, sex at birth, stimulant medication status, and mean neuromelanin signal in the substantia nigra pars compacta.

| Effect | Estimate (*B*) | *SE* (*B*) | β* | $t_{165}$ | p |
|---|---|---|---|---|---|
| Intercept | -0.206 | 0.008 | - | -0.050 | 0.960 |
| Age | -0.007 | 0.008 | -0.070 | -0.908 | 0.365 |
| Sex at Birth (Female) | -0.005 | 0.008 | -0.047 | -0.597 | 0.552 |
| Stimulant Medication | 0.0004 | 0.008 | 0.004 | 0.048 | 0.961 |
| SNc NM Signal | -0.015 | 0.008 | -0.144 | -1.844 | 0.067 |

*Note.* N = 165, Error degrees of freedom = 160, R-squared = 0.030, Adjusted R-squared = 0.005, F-statistic versus constant model = 1.21, p-value = 0.307.

Model coefficients were estimated using robust regression with Huber weighting.

*Abbreviations*: SE, standard error; β*, standardized regression coefficient; SNc, substantia nigra pars compacta; NM, neuromelanin.

**Table S10.** Regression models predicting within-network resting-state functional connectivity in each of the default mode network, frontoparietal network, and cingulo-opercular network from the following predictors: intercept, age, sex at birth, stimulant medication status, and mean neuromelanin signal in the ventral tegmental area.

| Y = Default Mode Network Within-Network RSFC[a] | | | | | |
|---|---|---|---|---|---|
| **Effect** | **Estimate (*B*)** | ***SE* (*B*)** | **β*** | **$t_{165}$** | **p** |
| Intercept | 0.313 | 0.005 | - | 0.055 | 0.956 |
| Age | 0.006 | 0.005 | 0.079 | 1.045 | 0.298 |
| Sex at Birth (Female) | 0.011 | 0.005 | 0.162 | 2.114 | 0.036 |
| Stimulant Medication | -0.0004 | 0.005 | -0.006 | -0.076 | 0.939 |
| VTA NM Signal | 0.016 | 0.005 | 0.224 | 2.928 | 0.004 |
| **Y = Frontoparietal Network Within-Network RSFC[b]** | | | | | |
| **Effect** | **Estimate (*B*)** | ***SE* (*B*)** | **β*** | **$t_{165}$** | **p** |
| Intercept | 0.198 | 0.003 | - | -0.227 | 0.824 |
| Age | -0.004 | 0.003 | -0.098 | -1.263 | 0.209 |
| Sex at Birth (Female) | 0.008 | 0.003 | 0.181 | 2.322 | 0.022 |
| Stimulant Medication | -0.002 | 0.003 | -0.036 | -0.465 | 0.642 |
| VTA NM Signal | -0.005 | 0.003 | -0.106 | -1.361 | 0.175 |
| **Y = Cingulo-opercular Network Within-Network RSFC[c]** | | | | | |
| **Effect** | **Estimate (*B*)** | ***SE* (*B*)** | **β*** | **$t_{165}$** | **p** |
| Intercept | 0.309 | 0.005 | - | -0.341 | 0.734 |
| Age | 0.002 | 0.005 | 0.039 | 0.484 | 0.629 |
| Sex at Birth (Female) | -0.0001 | 0.005 | -0.002 | -0.023 | 0.982 |
| Stimulant Medication | -0.001 | 0.005 | -0.023 | -0.287 | 0.775 |
| VTA NM Signal | 0.001 | 0.005 | 0.020 | 0.246 | 0.806 |

[a]N = 165, Error degrees of freedom = 160, R-squared = 0.088, Adjusted R-squared = 0.065, F-statistic versus constant model = 3.84, p-value = 0.005.

[b]N = 165, Error degrees of freedom = 160, R-squared = 0.052, Adjusted R-squared = 0.028, F-statistic versus constant model = 2.18, p-value = 0.074.

[c]N = 165, Error degrees of freedom = 160, R-squared = 0.003, Adjusted R-squared = -0.022, F-statistic versus constant model = 0.117, p-value = 0.976.

Model coefficients were estimated using robust regression with Huber weighting.

*Abbreviations*: SE, standard error; β*, standardized regression coefficient; VTA, ventral tegmental area; NM, neuromelanin.

**Table S11.** Regression models predicting network-specific system segregation in each of the default mode network, frontoparietal network, and cingulo-opercular network, as well as global system segregation, from the following predictors: intercept, age, sex at birth, stimulant medication status, and mean neuromelanin signal in the ventral tegmental area.

| **Y = Default Mode Network-specific System Segregation[a]** | | | | | |
|---|---|---|---|---|---|
| **Effect** | **Estimate (*B*)** | ***SE* (*B*)** | **β*** | **$t_{165}$** | **p** |
| Intercept | 1.253 | 0.009 | - | 0.854 | 0.394 |
| Age | 0.010 | 0.009 | 0.077 | 1.099 | 0.273 |
| Sex at Birth (Female) | -0.021 | 0.009 | 0.172 | -2.438 | 0.016 |
| Stimulant Medication | -0.005 | 0.009 | -0.039 | -0.556 | 0.579 |
| VTA NM Signal | 0.004 | 0.009 | 0.031 | 0.437 | 0.663 |
| **Y = Frontoparietal Network-specific System Segregation[b]** | | | | | |
| **Effect** | **Estimate (*B*)** | ***SE* (*B*)** | **β*** | **$t_{165}$** | **p** |
| Intercept | 0.941 | 0.010 | - | 0.810 | 0.421 |
| Age | 0.010 | 0.010 | 0.073 | 0.996 | 0.321 |
| Sex at Birth (Female) | -0.014 | 0.010 | -0.106 | -1.435 | 0.153 |
| Stimulant Medication | -0.008 | 0.010 | -0.062 | -0.845 | 0.399 |
| VTA NM Signal | 0.006 | 0.010 | 0.043 | 0.584 | 0.560 |
| **Y = Cingulo-opercular Network-specific System Segregation[c]** | | | | | |
| **Effect** | **Estimate (*B*)** | ***SE* (*B*)** | **β*** | **$t_{165}$** | **p** |
| Intercept | 1.389 | 0.010 | - | 0.879 | 0.381 |
| Age | 0.020 | 0.010 | 0.143 | 2.035 | 0.043 |
| Sex at Birth (Female) | 0.033 | 0.010 | 0.242 | 3.431 | 0.0008 |
| Stimulant Medication | -0.003 | 0.010 | -0.025 | -0.361 | 0.718 |
| VTA NM Signal | 0.019 | 0.010 | 0.137 | 1.936 | 0.055 |
| **Y = Global System Segregation[d]** | | | | | |
| **Effect** | **Estimate (*B*)** | ***SE* (*B*)** | **β*** | **$t_{165}$** | **p** |
| Intercept | 1.228 | 0.008 | - | 1.644 | 0.102 |
| Age | 0.012 | 0.008 | 0.110 | 1.578 | 0.117 |
| Sex at Birth (Female) | -0.003 | 0.008 | -0.029 | -0.416 | 0.678 |
| Stimulant Medication | -0.004 | 0.008 | -0.036 | -0.525 | 0.6006 |
| VTA NM Signal | 0.010 | 0.008 | 0.095 | 1.35 | 0.179 |

[a]N = 165, Error degrees of freedom = 160, R-squared = 0.050, Adjusted R-squared = 0.026, F-statistic versus constant model = 2.11, p-value = 0.082.
[b]N = 165, Error degrees of freedom = 160, R-squared = 0.028, Adjusted R-squared = 0.003, F-statistic versus constant model = 1.14, p-value = 0.340.
[c]N = 165, Error degrees of freedom = 160, R-squared = 0.117, Adjusted R-squared = 0.095, F-statistic versus constant model = 5.28, p-value = 0.0005.
[d]N = 165, Error degrees of freedom = 160, R-squared = 0.042, Adjusted R-squared = 0.018, F-statistic versus constant model = 1.75, p-value = 0.141.
Model coefficients were estimated using robust regression with Huber weighting.
*Abbreviations*: SE, standard error; β*, standardized regression coefficient; VTA, ventral tegmental area; NM, neuromelanin.

**Table S12.** Labels, anatomical regions, and degree centrality for each Cole-Anticevic Brain Network Parcellation (CAB-NP) parcel identified as a node in the neuromelanin-specific subnetwork identified via network-based statistic.

| CAB-NP Parcel Label | Anatomical Region | Degree Centrality |
|---|---|---|
| Cingulo-Opercular-29_R-Ctx | Dorsal Anterior Cingulate Gyrus | 12 |
| Default-01_R-Ctx | Precuneus | 1 |
| Default-02_R-Ctx | Parieto-occipital Sulcus | 1 |
| Default-04_R-Ctx | Posterior Cingulate Gyrus | 2 |
| Default-05_R-Ctx | Posterior Cingulate Gyrus | 1 |
| Default-06_R-Ctx | Posterior Cingulate Gyrus | 5 |
| Default-07_R-Ctx | Ventral Anterior Cingulate Gyrus | 1 |
| Default-08_R-Ctx | Ventromedial Prefrontal Cortex | 2 |
| Default-13_R-Ctx | Dorsomedial Prefrontal Cortex | 5 |
| Default-16_R-Ctx | Ventromedial Prefrontal Cortex | 1 |
| Default-28_R-Ctx | Temporal Pole | 3 |
| Default-37_R-Ctx | Superior Temporal Sulcus | 2 |
| Default-38_L-Ctx | Precuneus | 2 |
| Default-39_L-Ctx | Parieto-occipital Sulcus | 2 |
| Default-40_L-Ctx | Posterior Cingulate Gyrus | 3 |
| Default-41_L-Ctx | Retrosplenial Cortex | 1 |
| Default-42_L-Ctx | Posterior Cingulate Gyrus | 3 |
| Default-43_L-Ctx | Posterior Cingulate Gyrus | 4 |
| Default-44_L-Ctx | Ventral Anterior Cingulate Gyrus | 4 |
| Default-47_L-Ctx | Ventromedial Prefrontal Cortex | 1 |
| Default-52_L-Ctx | Pre-supplementary Motor Area | 1 |
| Default-57_L-Ctx | Ventromedial Prefrontal Cortex | 1 |
| Default-60_L-Ctx | Inferior Frontal Gyrus | 1 |
| Default-67_L-Ctx | Superior Temporal Sulcus | 2 |
| Default-69_L-Ctx | Inferior Parietal Lobe | 1 |
| Default-71_L-Ctx | Inferior Temporal Sulcus | 4 |
| Default-72_L-Ctx | Posterior Cingulate Gyrus | 3 |
| Default-76_L-Ctx | Superior Temporal Sulcus | 2 |
| Frontoparietal-05_R-Ctx | Dorsomedial Prefrontal Cortex | 1 |

**Table S13.** Labels, anatomical regions, and degree centrality for each Cole-Anticevic Brain Network Parcellation (CAB-NP) parcel identified as a node in both the neuromelanin-specific and hallucination-specific subnetworks identified via network-based statistic.

| CAB-NP Parcel Label | Anatomical Region | Degree Centrality: Hallucination | Degree Centrality: Neuromelanin |
|---|---|---|---|
| Default-04_R-Ctx | Posterior Cingulate Gyrus | 1 | 2 |
| Default-06_R-Ctx | Posterior Cingulate Gyrus | 1 | 5 |
| Default-07_R-Ctx | Ventral Anterior Cingulate Gyrus | 3 | 1 |
| Default-40_L-Ctx | Posterior Cingulate Gyrus | 1 | 3 |
| Default-44_L-Ctx | Ventral Anterior Cingulate Gyrus | 3 | 4 |
| Default-47_L-Ctx | Ventromedial Prefrontal Cortex | 8 | 1 |
| Default-57_L-Ctx | Ventromedial Prefrontal Cortex | 1 | 1 |
| Default-69_L-Ctx | Inferior Parietal Lobe | 1 | 1 |
